# Unveiling the Genetic Landscape of Basal Ganglia: Implications for Common Brain Disorders

**DOI:** 10.1101/2023.07.26.23293206

**Authors:** Shahram Bahrami, Kaja Nordengen, Jaroslav Rokicki, Alexey A. Shadrin, Zillur Rahman, Olav B. Smeland, Piotr Pawel Jaholkowski, Nadine Parker, Pravesh Parekh, Kevin S. O’Connell, Torbjørn Elvsåshagen, Mathias Toft, Srdjan Djurovic, Anders M. Dale, Lars T. Westlye, Tobias Kaufmann, Ole A. Andreassen

**Affiliations:** Institute of Clinical Medicine, University of Oslo, Oslo, Norway; KG Jebsen Centre for Neurodevelopmental disorders, University of Oslo, Oslo, Norway; Department of Neurology, Oslo University Hospital, Oslo, Norway; Centre of Research and Education in Forensic Psychiatry, Oslo University Hospital, Oslo, Norway; Department of Behavioral Medicine, Institute of Basic Medical Sciences, University of Oslo, Oslo, Norway; Department of Medical Genetics, Oslo University Hospital, Oslo, Norway; Multimodal Imaging Laboratory, University of California San Diego, La Jolla, USA; Department of Psychiatry, University of California, San Diego, La Jolla, USA; Department of Neurosciences, University of California San Diego, La Jolla, USA; Department of Radiology, University of California, San Diego, La Jolla, USA; Department of Psychology, Faculty of Social Sciences, University of Oslo, Norway; Department of Psychiatry and Psychotherapy, Tübingen Center for Mental Health, University of Tübingen, Germany; German Center for Mental Health (DZPG), Germany; Department of Psychiatry, Oslo University Hospital, Oslo, Norway

## Abstract

The basal ganglia are subcortical brain structures involved in motor control, cognition, and emotion regulation. We conducted a multivariate genome-wide association analysis (GWAS) to explore the genetic architecture of basal ganglia volumes using brain scans obtained from 34,794 European individuals with replication in 5,236 non-Europeans. We identified 72 genetic loci associated with basal ganglia volumes with a replication rate of 87.5%, revealing a distributed genetic architecture across basal ganglia structures. Of the 72 loci, 51 are novel. Of these, *APOE*, *NBR1* and *HLAA*, are all exonic and among the novel loci. Furthermore, we examined the genetic overlap between basal ganglia volumes and several neurological and psychiatric disorders. The most prominent overlap was seen with Parkinson’s disease, schizophrenia and migraine. *HP* and *TMEM161B* showed overlap between basal ganglia and Parkinson’s disease, but also three different psyciatric or nevrodevelopmental disorder(s), demonstrating important shared biology between brain disorders. Functional analyses implicated neurogenesis, neuron differentiation and development in basal ganglia volumes. These results enhance our understanding of the genetic architecture and molecular associations of basal ganglia structure and their role in brain disorders.

## Introduction

The basal ganglia are a group of interconnected subcortical nuclei deep in the brain^1^. The major parts of the basal ganglia are located in the cerebrum and include the caudate nucleus, putamen, globus pallidus and the accumbens area^2, 3^. Substantia nigra in the midbrain and the subthalamic nucleus in diencephalon may be regarded as associated structures. Although the basal ganglia comprise physically distinct entities, they exhibit a robust functional cohesion due to their intricate interconnections with numerous pathways.

The basal ganglia integrate and modulate cortical information, and are involved in motor^4^, cognitive^5^ and limbic functions^3, 6^. Specifically, they are involved in motor functions through initiation, execution, and coordination of movements^7^, action selection^5^ and the inhibition of unwanted or competing motor responses. The basal ganglia are also involved in learning and execution of procedural memory and habits, allowing actions to become automatic and efficient over time ^8, 9^. Beyond motor control, the basal ganglia play important roles in cognitive functions like decision-making^5^, the shifting of attention, updating information, and adapting behavior to changing circumstances^5, 10–12^, in addition to the anticipation, evaluation, and processing of reward, which shapes behavior, decision-making^13^ and motivation^14^. Finally, the basal ganglia are involved in emotional processing and the regulation of affective states^3,6^, and contribute to the integration of this emotional information in decision-making processes^3,15^.

Likely due to their widespread connections to other parts of the brain, the basal ganglia have been implicated in several important brain diseases, ranging from neurodegenerative to psychiatric and neurodevelopmental conditions. The most well-known disorder related to the basal ganglia is Parkinson’s disease (PD), where loss of dopaminergic projections from the substantia nigra to the basal ganglia lead to progressive motor symptoms. In Alzheimer’s disease (ALZ), evidence suggests that tau- and amyloid aggregation can affect the basal ganglia and contribute to cognitive and motor impairments^16^. Through pain processing and modulation, basal ganglia are also involved in the pathophysiology of the primary headache disorder migraine (MIG). Dysfunction within the basal ganglia circuits may contribute to stereotyped/repetitive movements, reduced attentional control^11, 12^ and social and emotional processing^6^ often observed in individuals with autism spectrum disorder (ASD)^17, 18^. Through its role in regulating motor hyperactivity^19, 20^, reward processing^21–23^ and inhibitory control^24, 25^, the basal ganglia is also central for the pathophysiology of attention-deficit/hyperactivity disorder (ADHD)^24–26^. Dysregulation of dopamine neurotransmission in the mesocorticolimbic pathway involving the basal ganglia has been implicated in ADHD^27–29^, but also related to hallucinations and delusions in schizophrenia (SCZ)^30, 31^, while cortico-striatal circuitry abnormalities may contribute to the cognitive impairments observed in SCZ^32, 33^. The basal ganglia also play a role in mood regulation and emotional processing^34, 35^, and basal ganglia functions have been implicated in both bipolar disorder (BIP)^35^ and major depressive disorder (MDD) ^34, 36^.

Despite the significance of the basal ganglia in motor control, decision-making, reward processing, and emotional regulation^3^, as well as in pathogenesis of neurological and psychiatric disorders, the genetic factors influencing their volumes have not been comprehensively characterized in previous research. Studies of basal ganglia volumes have in many circumstances showed inconsistent results. For MIG^37^, PD^38^, MDD^39^, ASD^12^, and ADHD^40, 41^ some studies report reduced basal ganglia volumes, while others report no significant differences ^42, 43^, or even striatal enlargement ^44–48^.

While the functional aspects of the basal ganglia have been extensively studied, the genetic architecture underlying their structural characteristics remains largely unexplored. There are, however, genetic studies on volumetrics of subcortical structures which includes basal ganglia nuclei, ^49, 50^ without focusing on the nuclei as a functional unit. Comprehensive understanding of the genetic mechanisms shaping basal ganglia volumes is lacking. This study represents the first multivariate genome wide association study (GWAS) on the basal ganglia volumes, as a representation of this as one functional unit. Understanding the genetic basis of basal ganglia volumes as a whole can provide insights into the pathogenesis and etiology of basal ganglia related brain disorders, which can form the basis for future development of targeted therapeutic interventions.

## Results

All basal ganglia volumes showed significant SNP-heritability (Figure 1A), with highest estimates for the caudate (h2=0.34, se=0.025) and the whole basal ganglia (h2 = 0.31, se=0.026). Figure 1B shows a correlation matrix of basal ganglia volumes, with phenotypic correlation shown in upper-left section and genetic correlation using the univariate GWAS summary statistics shown in the lower-right section. As expected, genetic correlations mapped phenotypic correlations (Supplementary Table 1). The overall lowest genetic correlations was between pallidum and accumbens (rg= 0.1153, se= 0.056), which are also the structures with lowest estimated heritability.

**Figure 1.**
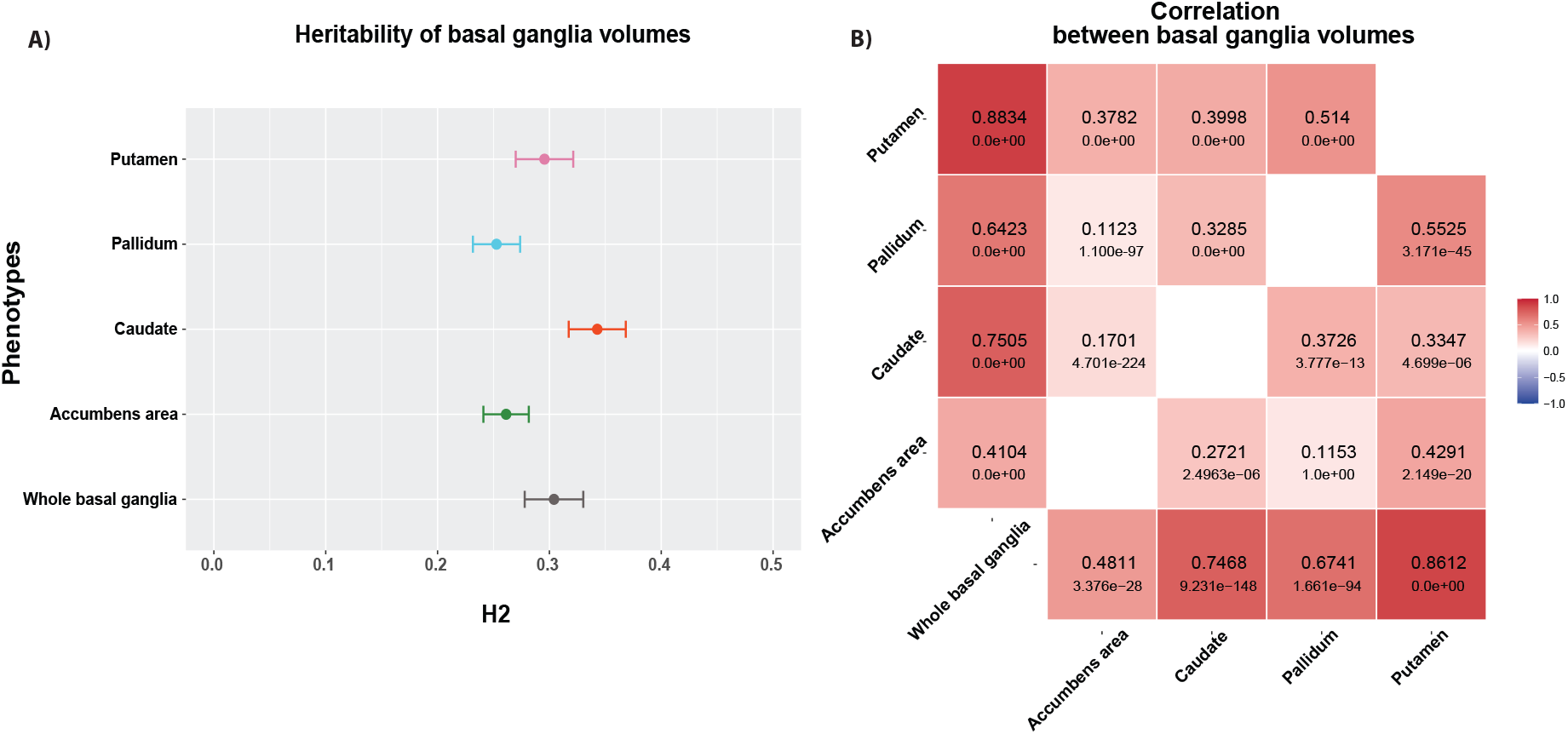
The univariate signatures of basal ganglia volumes indicate high heritability and significant phenotypic and genetic correlations. **A**. Heritability of basal ganglia volumes, where the numbers depict SNP-heritability estimates (h2). Error bars reflect standard error. **B.** LD-score regression-based genetic correlations (in lower-right section) and phenotypic correlation (in upper-left section) between each pair of regions, using the univariate GWAS summary statistics.

### Multivariate GWAS reveals 72 genetic loci associated with basal ganglia

A multivariate GWAS deployed using the MOSTest framework^54^ identified 72 significant independent loci, including 51 novel loci (Supplementary Table 2). We thresholded based on genome-wide significance (P < 5e−8) and identified a total of 12305 candidate SNPs, 294 independent significant SNPs and 89 lead SNPs across structures located in the 72 genomic loci, using the FUMA platform^58^ (Supplementary Table 3). Figure 2 illustrates the results of multivariate and univariate GWAS, where the upper part of the Miami plot shows the multivariate polygenic architecture across basal ganglia volumes. A distributed genetic architecture throughout the basal ganglia structure is supported by the higher multivariate statistics in comparison with the univariate statistics for various basal ganglia volumes in the most of the identified loci (Figure 1), which is further confirmed by genetic correlation analysis of the individual volumes (Supplementary Table 1).

**Figure 2.**
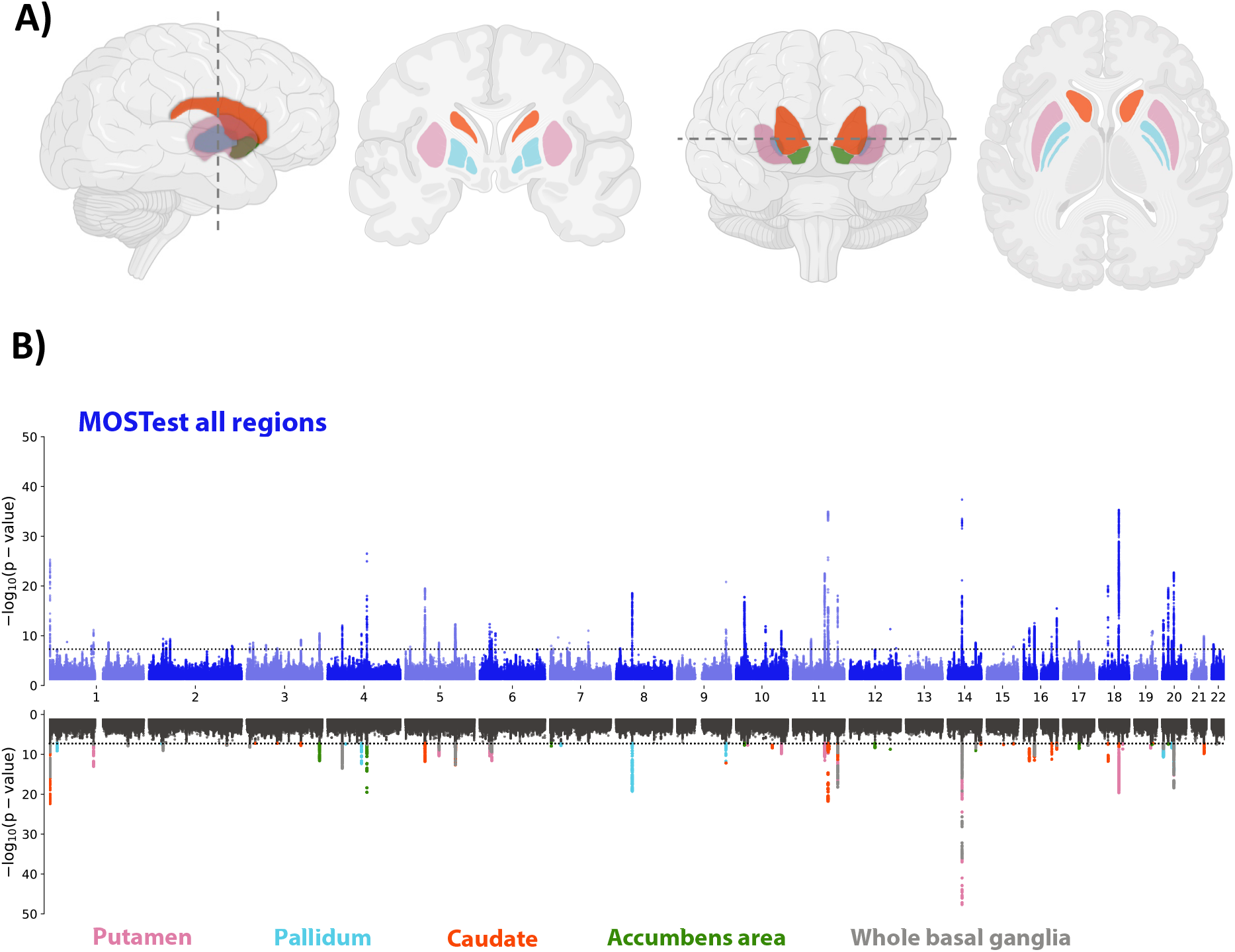
The genetic architecture of basal ganglia. **A.** Schematic illustration of the basal ganglia regions, comprising the histologically distinguishable subfields of putamen, pallidum, caudate and accumbens. **B.** The upper part of the Miami plot illustrates the −log10(*P*) statistic from the multivariate GWAS across the entire basal ganglia, with 72 significant loci. For comparison, the lower part depicts for each of the 72 unique loci the corresponding −log10(*P*) statistics from univariate GWASs of single subregions (one colour per subregion, black indicates non-significant SNPs, *p*-values are two-tailed), supporting a distributed genetic architecture across the basal ganglia structure.

Although the strongest associations among the 72 significant independent loci that were identified in the multivariate framework are also present in the results of univariate analysis, a considerable proportion of these loci demonstrated higher effects that were not significant at the genome-wide level. Leveraging these distributed effects across the various subregions, the multivariate approach resulted in enhanced discovery. Q-Q plots from MOSTest analysis (Supplementary Figure 1) including one from permutation testing, showed successive validity of the multivariate test statistic. The findings were supported by a multivariate replication study that produced the same effect direction for 87.5% of the lead SNPs in a separate sample of 5220 non-white people (Supplementary Figure 2).

### Functional annotation, gene mapping and genetic analyses

We functionally annotated SNPs associated with basal ganglia volumes that were in LD (r2 ≥ 0.6) with one of the independent significant SNPs with P < 5e−8 in the discovery sample using FUMA v1.4.1^58^. A majority of these SNPs were intronic (53.5%) or intergenic (29.1%) and 1.1% were exonic (Supplementary Figure 3A and Supplementary Table 3). Supplementary Figure 3B provides information for functional SNP categories for the basal ganglia volume. About 81.8% of the SNPs had a minimum chromatin state of 1–7, thus suggesting they were in open chromatin regions (Supplementary Figure 3C)^60, 80^. Three of the lead SNPs were exonic and combined annotation-dependent depletion (CADD) scores of those SNPs were 23.1 (rs13107325), 18.4 (rs2070835) and 12.64 (rs429358), thus indicating deleterious protein effects^59^ (Supplementary Table 2). rs13107325 and rs429358 are located in *SLC39A8* and *APOE*, respectively, and have previously been associated with SCZ, PD and Alz^81–84^.

Genome-wide gene-based association analyses (GWGAS; P < 2.622e-6, i.e., 0.05/ 19073 genes) using MAGMA v1.08^61^ detected 149 unique genes across the basal ganglia (Supplementary Table 4). Supplementary Figure 4 provides Manhattan and Q–Q plots for the GWGAS. Gene-set analyses using MAGMA identified significant Gene Ontology sets for neurogenesis, neuron differentiation and development (Supplementary Table 5).

### Open Target Genetics

We additionally used Open Target Genetics to identify the target genes for each lead SNP of the 72 loci implied for the basal ganglia volumes (Figure 3 & Supplementary Table 2).

**Figure 3.**
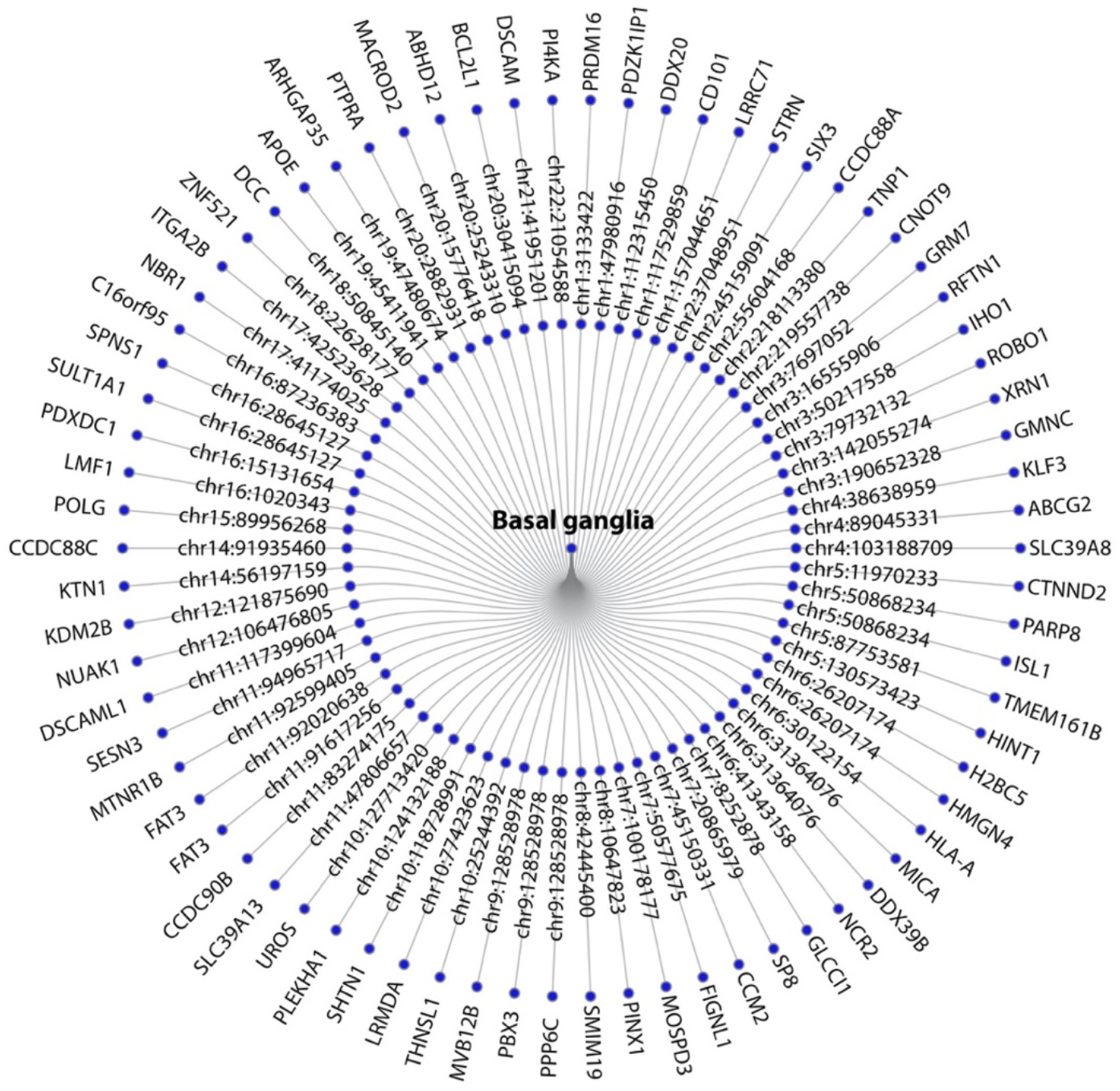
Mapped genes to the significant loci. Gene mapping of the 72 loci associated with the basal ganglia implied 73 genes by Open target.

We used the 73 uniquely mapped genes from Open Targets for gene set analysis, pathway analysis and differentially expressed genes (DEG) analysis. GO gene-set analysis for the target genes of the significant loci revealed 16 significantly associated biological processes, including ‘GO_NEURON_DIFFERENTIATION’, ‘GO_NEUROGENESIS’ and ‘GO_CELL_PART_MORPHOGENESIS’ (Supplementary Table 6). These genes were also significantly associated with four cellular component gene-sets, including ‘GO_NEURON_PROJECTION’ and ‘GO_SYNAPSE’ (Supplementary Table 6). There were 19 pathways significantly overrepresented among the target genes of the significant loci with ‘the DSCAM interactions’, ‘Axon guidance’, ‘Nervous system development’ and ‘Netrin-1 signaling’as the most significant (Supplementary Table 7).

We also tested the tissue specificity using the DEG sets defined for the target genes of the identified loci. The results show that these target genes are significantly expressed in the brain and specifically in basal ganglia (Supplementary Figure 5 & 6).

In addition, we found enrichment for the mapped genes in the lymphatic system, nervous systems, and sensory systems (Supplementary Figure 7A); the top enriched cell types were mostly related to the immune and nervous systems (natural killer cells, T cells, dendritic cells, bipolar neurons, inhibitory neurons, and endothelial cells) (Supplementary Figure 7B). We also showed that the mapped genes expressed in the brain have cell-type-specific expression patterns in the main cell types of the human cerebral cortex (Supplementary Figure 8).

We determined protein–protein and co-expression networks for the mapped genes. The genes have 14.17 and 16.41 physical interaction and gene co-expression, respectively (Supplementary Figure 9). A gene drug interaction analysis shows that 13 out of 73 genes have interaction with 130 drugs (Supplementary Table 8), including antipsychotic (risperidone, chlorpromazine) and antiepileptic/mood stabilizing drugs (lamotrigine, carbamazepine).

### Genetic overlap between the basal ganglia and common brain disorders

To further examine the polygenic architecture of basal ganglia volumes and the potential genetic overlap between basal ganglia and common brain disorders, we used GWAS summary statistics for ADHD, ASD, BIP, MDD, SCZ, Alz, MIG, and PD. Genetic correlations of the disorders with individual basal ganglia subregions revealed only a significant association with PD after Bonferroni correction (Supplementary Figure 10 & Supplementary Table 9). Conditional Q–Q plots conditioning the multivariate statistic of basal ganglia on the disorders and vice versa clearly demonstrated a pattern of pleiotropic enrichment in both directions (Supplementary Figure 11). Conjunctional FDR analysis allowed us to test for shared loci between the basal ganglia and each of the disorders. We identified 3 loci significantly overlapping with ADHD, 2 loci with ASD, 20 with BIP, 83 with SCZ, 15 with MDD, 33 with MIG, 21 with Alz and 28 with PD. (Figure 4 & Supplementary Table 10-17)

**Figure 4.**
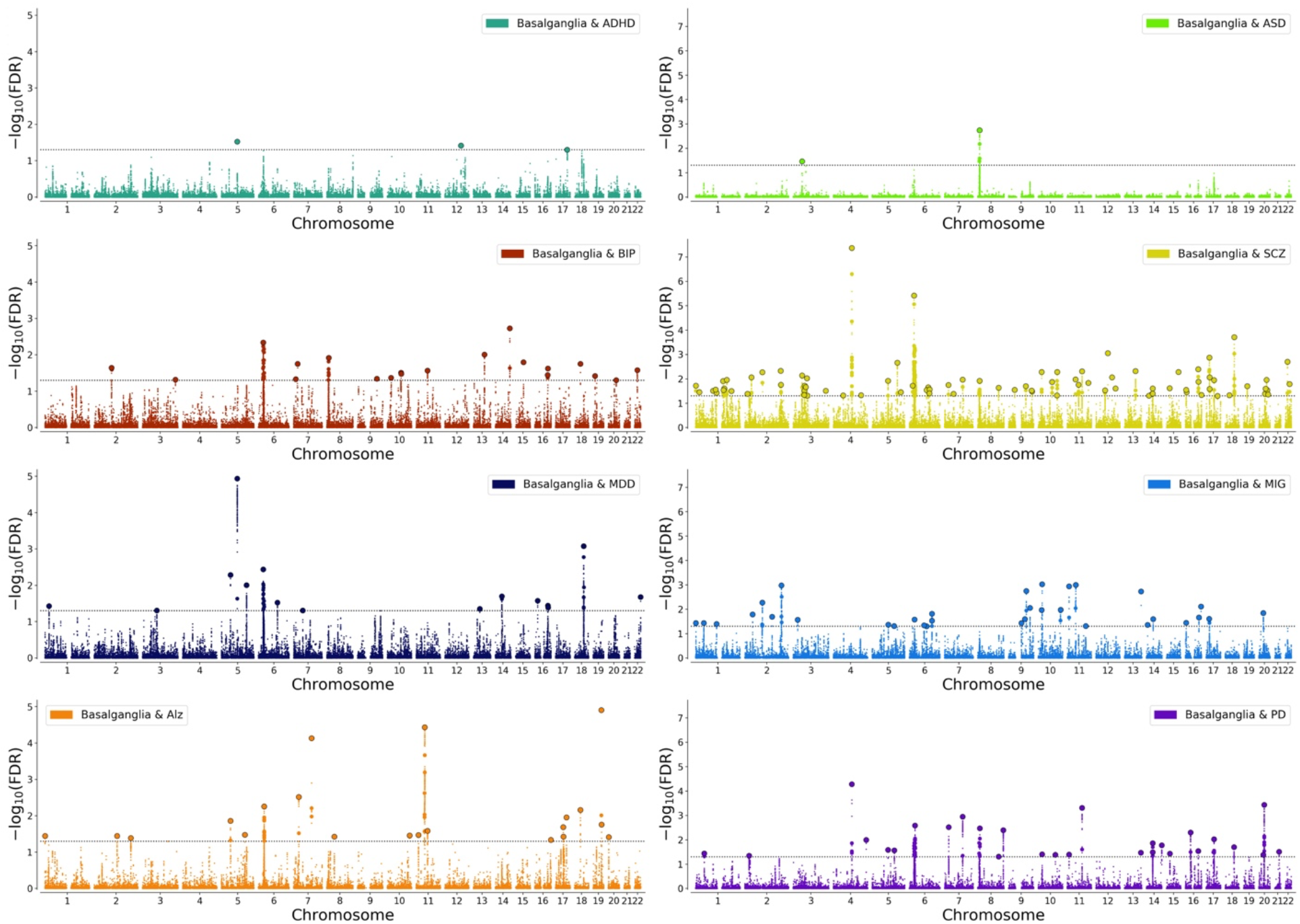
Genetic overlap between basal ganglia and common brain disorders. Conjunctional FDR Manhattan plots, showing the –log10 transformed conjunctional FDR values for each SNP on the y-axis and chromosomal positions along the x-axis. The dotted horizontal line represents the threshold for significant shared associations (conjFDR < 0.05). Independent lead SNPs are encircled in black.

### Gene mapping and overlapping shared significant loci and genes

A full list of loci overlapping between basal ganglia and the eight disorders is provided in Supplementary Tables 10-17. Figure 5 illustrates the number of significantly shared loci and genes across each combination of disorders. First, we identified significant shared loci according to the FUMA protocol^58^ for each conjFDR analyses. Across all phenotypes, we grouped physically overlapping loci, resulting in a total number of 159 distinct loci including 103 loci that were associated with psychiatric disorders and 73 loci associated with neurological disorders. In total, 17 loci were shared between neurological and psychiatric disorders.

**Figure 5.**
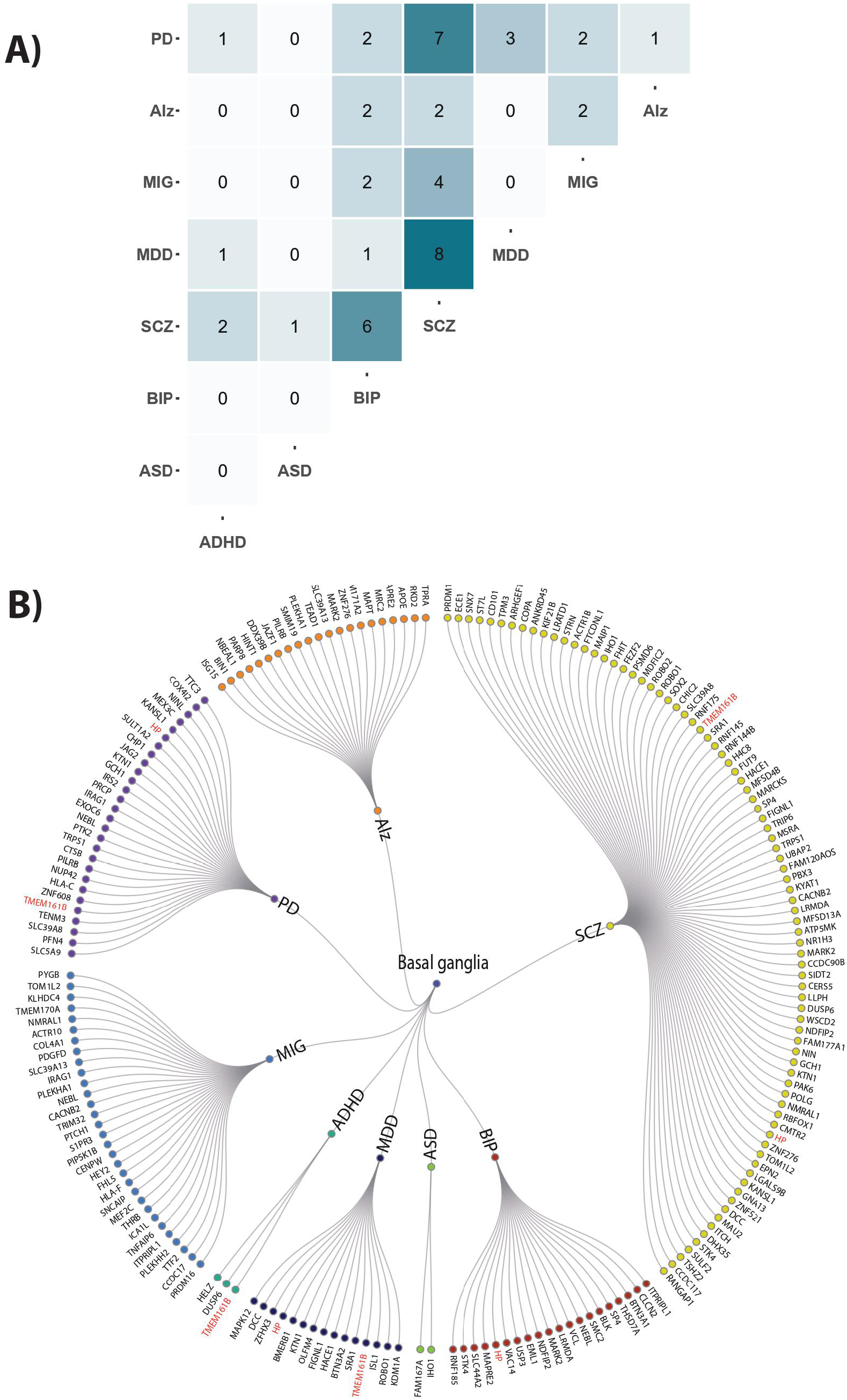
Various genes mapped from the conjunctional FDR analysis were implied to overlap between basal ganglia and multiple disorders. **A.** The figure shows the total number of genes overlapping for each combination of disorders. For example, 8 of the genes overlapping between basal ganglia and SCZ were also found to overlap between basal ganglia and MDD. **B.** All mapped genes to overlapped loci between basal ganglia and disorders and genes that were implied for more than 4 disorders (shown in red). Genes *TMEM161B* and *HP* were mapped for four disorders. ASD autism spectrum disorder, ADHD attention deficit hyperactivity disorder, SCZ schizophrenia, BIP bipolar disorder, MIG migraine, MDD, major depression, PD Parkinson’s disease, Alz Alzheimer’s disease.

Next, we mapped each of these shared loci to significantly associated protein-coding genes using Open target mapping (Supplementary Table 10-17). Overall, the extent of gene pleiotropy was similar to that observed for loci. A total of 165 distinct genes were identified across all phenotypes. Of these, 106 and 75 genes were associated with psychiatric and neurological disorders, respectively, of which 16 were overlapping (Table 1).

**Table 1.**
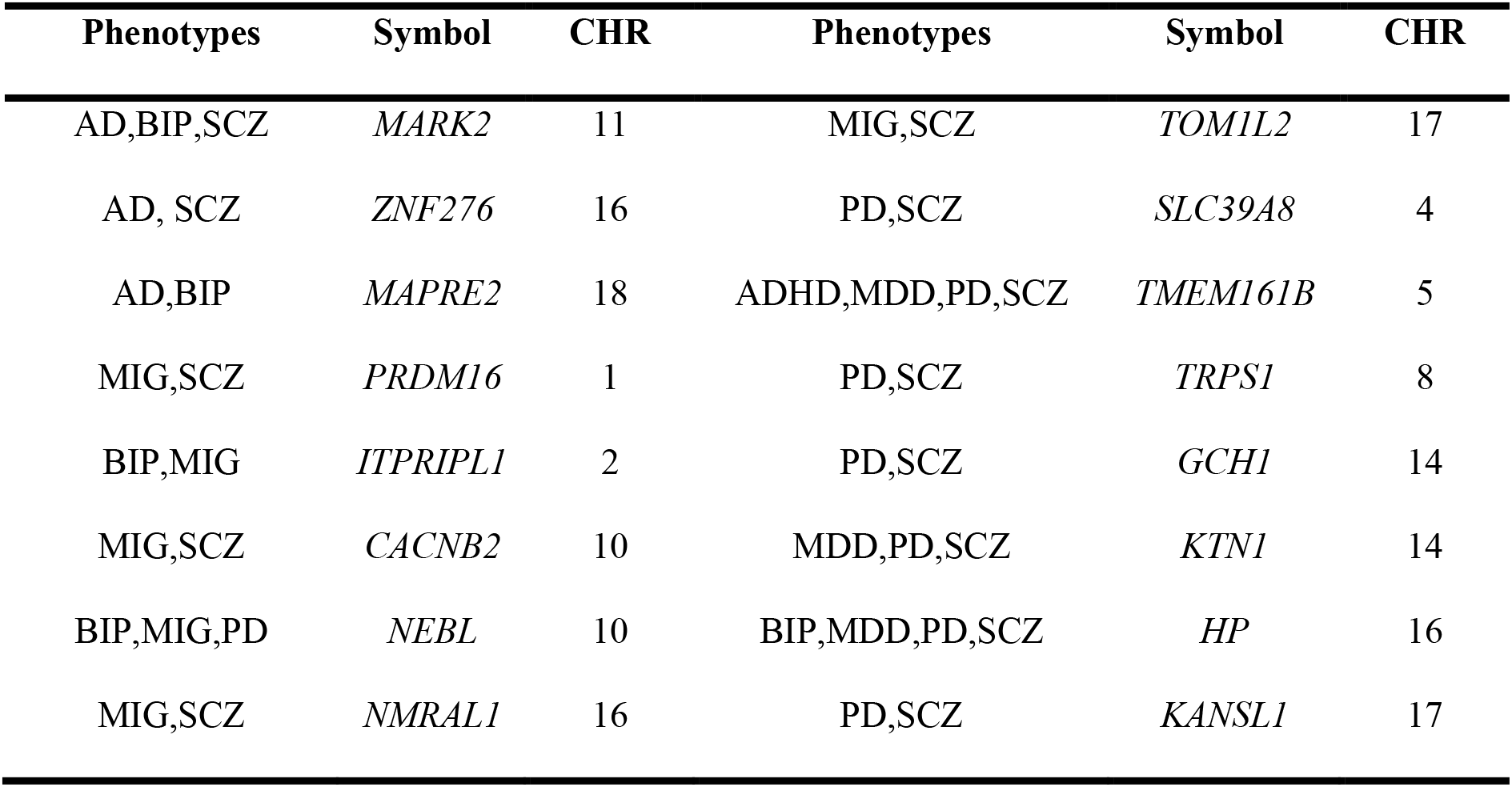
Overlapping genes between basal ganglia, psychiatric and neurological disorders.

We also examined genes that were implicated in multiple disorders. By far the strongest overlap was found between SCZ and MDD, where 8 of the genes overlapping between basal ganglia and SCZ were also found to overlap between basal ganglia and MDD (Figure 5 A). We also found large overlap between other combinations of disorders, such as SCZ and PD (7 genes), and SCZ and BIP (6 genes) (Figure 5 A).

Some of the genes were implicated in more than two disorders. Figure 5 B shows all genes overlapping between the basal ganglia volumes and the 8 disorders. The most frequently mapped genes were the *HP* and *TMEM161B* genes, which overlapped between basal ganglia and at least four disorders. (Figure 5 B).

## Discussion

In summary, our multivariate GWAS of basal ganglia volumes uncovered numerous genomic loci that were not previously identified, indicating that the effects on basal ganglia structures are distributed across basal ganglia structures in line with their function as one unit. The mapped genes have pivotal roles in neurotransmission, neuronal differentiation and synaptogenesis, underscoring their significance in shaping neural circuitry and facilitating proper brain development. The overlapping genetic architecture and shared genes between basal ganglia and common neurological and psychiatric disorders may pinpoint potential disease-independent drug targets.

The distributed genetic architecture across the basal ganglia, here revealed through 72 associated loci implicating 73 target genes, pointed at early developmental pathways shared between neurological and psychiatric disorders, dominated by embryogenic brain development, with neuronal projection guidance, neurogenesis and neuronal differentiation for genes mapped to this first ever basal ganglia GWAS. The precise embryonic organization of the basal ganglia exert a profound influence on its function in both health and disease, including its volumetric attributes, being centrally positioned deep in the brain with vital projections to the frontal cortex and limbic structures ^3,15^. Functional analyses further implicated locomotor behavior, which encompass voluntary movements such as walking and coordination of the limbs. The basal ganglia and its connections contribute to the regulation and execution of locomotor activity. Understanding the genetic factors underlying basal ganglia involvement in locomotion is important for deciphering the complexities of motor control, and can potentially guide the development of targeted therapeutic strategies for movement disorders.

Our findings align with and expand upon earlier reports from univariate analyses of studies including basal ganglia volumes together with other subcortical structures. Hibar et al 2015^49^ discovered 15 loci associated with striatal structures and Satizabel et al 2019^50^ discovered 48 loci significantly associated with subcortical structures, of which 21 was replicated in our multivariate basal ganglia GWAS. Of the 72 basal ganglia associated loci unveiled in this study, 51 loci were novel. Our two most significant loci associated with the basal ganglia have been consistently replicated from these previous univariate GWASs on subcortical volumes. One of these loci is located at the 11q14-3 region near the *FAT3* gene, which plays a crucial role in neuronal morphogenesis and cell migration as a conserved cellular adhesion molecule^85^. The second locus is situated at the 14q22 region near the *KTN1* gene, which encodes a kinesin-binding protein involved in the transport of cellular components along microtubules ^86^. Earlier univariate GWASs have previously found associations of *FAT3* to both caudate nucleus^49 50^ and putamen ^50^, and *KTN1* to putamen ^49 50^, in addition to accumbens, caudate nucleus and the globus pallidus ^50^. The wide range of associations with basal ganglia volumes contributes to the unquestionable significance of these two loci in our multivariate GWAS analysis. Additional findings that provide support for previous univariate GWAS results include our 208 different candidate SNPs are annotated to the 8p11.21 region associated with the present basal ganglia GWAS (see supplementary table 3), and previously seen associated to putamen in univariate GWAS^50^. 73 of these candidate SNPs is mapped to *SLC20A2* robustly linked to the familiar basal ganglia calcification ^87–89^, although the lead SNP is mapped to *SMIM19* gene, located in close proximity to *SLC20A2*.

The two undoubtedly most significant SNPs, and the above-mentioned SNPs mapped to SLC20A2 and SMIM19, are all in intergenic regions. However, four of the 72 basal ganglia associated loci have lead SNPs in the exonic regions of the genome (see suppl. figure 3 and supplementary table 2): *SLC39A8* on chr 4, *TRIM10* and *HLA-A* on chr 6, *VAT1* and *NBR1* on chr 17 and *APOE* on chr 19. *SLC39A8* has also previously been linked to basal ganglia^50^, is a transmembrane protein known to cotransport divalent cations with bicarbonate^90^, of which transport of manganese is suggested to be of greatest importance^91^, especially for dopaminergic projections in the basal ganglia^92^. The remaining three, however, are novel for this basal ganglia GWAS. rs3094134 on chr 6 located in the exon on *TRIM10* and mapped to *HLA-A*, both important for the immune functioning in the brain ^93–95^. For the first time, the Alz-linked *APOE* gene on chr 19, known to promote amyloid degradation^96^, are found genetically associated with basal ganglia. Several significant SNPs are annotated to the same region on chr 17 where rs8482 is in the exon of *NBR1*, encoding a autophagic adapter protein involved in Lewy body formation in PD^97^ and targeting ubiquitinated protein for degradation in general ^98^. The lead SNP in this locus, rs2070835, is located in the exonic region of *VAT1* (vesicle amine transport protein 1), which encodes a protein that plays a crucial role in the process of vesicular neurotransmitter transport^99^. Vat1 is primarily involved in the packaging and storage of monoamine neurotransmitters, such as dopamine, norepinephrine, and serotonin, all neurotransmitters distributed in the basal ganglia. *VAT1* and *NBR1* are located in close proximity to each other on 17q21, on each side of the tumor suppressor gene *BRCA1*. Mapped with Open Target *NBR1* is ranked first and *VAT1* second.

There is clear pleiotropic enrichment between basal ganglia volume and neurological (PD, AD, migraine), neurodevelopmental (ADHD, ASD) and psychiatric (BIP, MDD, SCZ) disorders, shown by Q-Q plots (Supplementary Figure 11), and many shared loci showed on Manhattan plots (figure 4) and in tables (supplementary tables 10-17). It is, however, only directly significant association between genetic risk of PD and genetic architecture of basal ganglia (Supplementary Figure 10 and supplementary table 9). The association between PD risk and the genetic pattern of basal ganglia volume, with all off the individual volumes pointing in the same direction, show a positive correlation. As a response to learning, there are commonly reported locally increased brain volume representing synaptogenesis and increased dendritic arborisation^100, 101^. Gene ontology analysis (supplementary table 7) support these mechanisms in the genetic architecture of basal ganglia volume, with gene ontology on cell part morphogenesis, neuron part and synapse, but details on cell structure changes representing the positive correlation between the genetic pattern of basal ganglia and PD need separate studies.

Our results suggest that there is large genetic pleiotropy between the basal ganglia and especially PD, SCZ and migraine, but also substantially to Alz, BIP and MDD, and less between ASD and ADHD. Out of more than 150 genes, only 16 genes were mapped to basal ganglia and both neurological and psychiatric disorders, indicating distinct genetic architecture underlying each of the disorder’s relationship with the basal ganglia. These 16 genes that show overlapping involvement in both neurological disorders and psychiatric diseases, demonstrate important shared biology between brain disorders (table 1). Notably, *TMEM161B* (transmembrane protein 161B) and *HP* (haptoglobin) is overlapping between basal ganglia and MDD, PD and SCZ, in addition to ADHD and BIP, respectively. Haptoglobin is involved in binding and transporting hemoglobin, and it plays a role in modulating the immune response and oxidative stress^102^. *TMEM161B* has been mapped to MDD in mouse models and humans^103, 104^ and has also been coupled to basal ganglia activation during reward processing^105^. Although functionally linked to basal ganglia, *TMEM161B* have previously only been linked to neocortex structurally, through gyrification in neocortical development^106^.

In summary, our findings indicate that the basal ganglia exhibit a polygenic architecture, spanning the putamen, caudate, pallidum, and accumbens, with high correlation between the basal ganglia subregions and a robust 72 loci when analyzed together, 51 of which is novel. The genetic overlap observed with different brain disorders highlights the interconnectedness of the basal ganglia in neurological (PD, ALZ, migraine) and psychiatric (SCZ, BIP, MDD) conditions, but less for neurodevelopmental conditions (ASD and ADHD). By recognizing the distributed nature of genetic effects on the brain, we can gain valuable insights into the mechanisms underlying brain disorders and pave the way for future advancements in this field. Moreover, our study reveals that some of these genetic findings align with known treatment targets, supporting the effectiveness of our approach. This may suggest shared underlying mechanisms and potential targets for therapeutic interventions. Further research is necessary to fully understand the complex relationship between the basal ganglia and these disorders and to develop more effective treatments.

## Data Availability

In this study we used brain imaging and genetics data from the UK Biobank [https://www.ukbiobank.ac.uk/], and GWAS summary statistics obtained from the Psychiatric Genomics Consortium [https://www.med.unc.edu/pgc/shared-methods/], 23andMe,inc. [https://www.23andme.com/], International headache genetics Consortium (IHGC) [http://www.headachegenetics.org/content/datasets-and-cohorts], the International Genomics of Alzheimer Project [https://ctg.cncr.nl/software/summary_statistics], and the International Parkinson Disease Genomics Consortium [https://pdgenetics.org/resources]. The latter included 23 and Me data, which was made available through 23andMe under an agreement with 23andMe that protects the privacy of the 23andMe participants [https://research.23andme.com/collaborate/#dataset-access/]. The summary statistics for basal ganglia derived in this study is available in our github repository [https://github.com/norment/open-science]. FUMA results are available online [https://fuma.ctglab.nl/browse/371].

## Acknowledgements

The authors were funded by Norwegian Health Association (SB: 22731, KN: 25598), the South-Eastern Norway Regional Health Authority (OAA: 2013-123, 2017-112, 2019-108), the Research Council of Norway (TK: 276082, 323961. OAA: 213837, 223273, 248778, 273291, 262656, 229129, 283798, 311993. LTW: 204966, 249795, 273345). LTW: 2014-097, 2015-073, 2016-083), Stiftelsen Kristian Gerhard Jebsen, the European Research Council (LTW: ERCStG 802998) and the Department of Neurology at Oslo University Hospital (KN). The funding bodies had no role in the analysis or interpretation of the data; the preparation, review or approval of the manuscript; nor in the decision to submit the manuscript for publication. This work was performed on the Tjeneste for Sensitive Data (TSD) facilities, owned by the University of Oslo, operated and developed by the TSD service group at the University of Oslo, IT-Department (USIT) and on resources provided by UNINETT Sigma2—the National Infrastructure for High Performance Computing and Data Storage in Norway. The research has been conducted using the UK Biobank Resource (access code 27412) and using summary statistics for various brain disorders that partly included 23andMe data. We would like to thank the research participants and employees of UK Biobank, the 23andMe, the Psychiatric Genomics Consortium, International headache genetics Consortium, the International Genomics of Alzheimer’s Project and International Parkinson Disease Genomics Consortium for contributing summary statistics for making this work possible.

## Author contributions

SB and KN conceived the study. SB and JR analysed the data. SB and KN interpreted the results and spearheaded the writing. SB drafted the online methods. All authors gave conceptual input on the methods and/or results and all authors contributed to and approved the final manuscript.

## Methods

### Sample and pre-processing of imaging and genetic data

We employed T1 weighted MRI data to investigate basal ganglia volumes. Our primary analysis was based on a dataset consisting of 34,794 genotyped individuals of white British descent from the UK Biobank (age range: 45-82 years, mean: 64.3 years, s.d.: 7.5 years, 52.3% females)^51^. To ensure the robustness of our findings, we also conducted a replication analysis using an independent dataset from UKB consisting of 5220 individuals with non-white ethnicity (age range: 45–81, mean: 62.9, s.d.: 7.4 years, 54.1% females).

We used Freesurfer v5.3 to extract the volumes of the accumbens, caudate, pallidum, and putamen^52^. For genetic analyses, we used the UK Biobank v3 imputed genetic data and followed the standard quality control procedures to remove SNPs with an imputation quality score of 0.5, a minor allele frequency <0.005, missingness in more than 10% of individuals, and failing the Hardy-Weinberg equilibrium tests at a P value <1e-9.

### Multivariate genome-wide association analysis

We calculated the average volume between the left and right hemispheres for each of the accumbens, caudate, pallidum, and putamen as well as the total basal ganglia volume. To address potential confounding factors, we employed neuroCombat^53^ to pre-residualize the data by accounting for age, age squared, sex, scanning site, Euler score (which serves as a proxy for image quality), total intracranial volume (ICV), and the 20 genetic principal components for each region comprising the basal ganglia. Furthermore, we conducted additional analyses, controlling for the total volume of the four basal ganglia volumes. This pre-residualization procedure was performed for both British and non-white ethnicity samples. We conducted a multivariate GWAS on pre-residualized basal ganglia volumes for our main analysis using Multivariate Omnibus Statistical Test (MOSTest)^54^, which is a powerful statistical tool used for joint genetic analysis of multiple traits in large-scale data. It aims to identify genetic variants associated with complex phenotypes influenced by multiple genetic factors with small effects. By analyzing the traits jointly, MOSTest takes a multivariate approach, considering multiple traits simultaneously and leveraging the genetic overlap across different regions and measures of the brain. This approach enhances the statistical power to detect associations by avoiding the stringent multiple comparison correction required in mass-univariate approaches. Van der Meer et al. (2020)^54^ provide detailed description of the method, while github.com/precimed/mostest provides information on the software implementation. We also conducted a univariate GWAS of the four basal ganglia volumes and whole basal ganglia (extracted from the univariate stream of MOSTest^54^) for comparison to conventional univariate techniques (GWAS). SNP-based heritability estimates for the basal ganglia volumes, as well as the genetic correlations between these volumes, were estimated using linkage disequilibrium (LD) score regression^55, 56^.

### Multivariate replication analysis

We used a multivariate replication approach developed to test if the multivariate pattern of genetic associations identified in discovery analysis is consistent between discovery and replication samples. This procedure generates a composite score from mass-univariate z-statistics for each locus identified in the multivariate analysis of the discovery sample, and tests for associations of the composite score with genotype in the replication sample (for mathematical formulation see Loughnan et al.^57^). We report the percent of loci replicating at P < 0.05 and the percent of loci showing the same effect direction.

### Functional annotation, gene-based association, gene-set, tissue, and pathway analysis

We submitted the summary statistic to the FUMA platform v1.4.1^58^ in order to identify independent genetic loci. We discovered independent significant SNPs at the statistical significance level P < 5e−8 using the 1000GPhase3 EUR as the reference panel. A portion of the independent significant SNPs in linkage equilibrium with one another at r^2^ < 0.1 were termed lead SNPs, and all SNPs at r^2^ < 0.6 with each other were considered independent significant SNPs. If two or more lead SNPs located within one LD block (in 250kb), we merged them into one genomic risk locus. Combined Annotation Dependent Depletion (CADD) scores, which forecast the deleteriousness of SNPs on protein structure/function^59^, RegulomeDB scores, which forecast regulatory functions^60^, and chromatin states, which illustrate the transcriptional/regulatory effects of chromatin states at the SNP locus, are all used by FUMA to annotate associated SNPs.

We conducted genome-wide gene-based association and gene-set analyses using MAGMA v.1.08^61^ in FUMA. MAGMA performs the gene-based association analysis and assigns p-values to individual genes based on the association between genetic variants within or near the gene and the trait of interest. This analysis is conducted using a multiple regression model, which improves statistical performance compared to single-marker analysis^61^. Furthermore, the gene-set analysis using MAGMA enables the assessment of groups of genes that share common biological functions or pathways, which provides insights into the functional and biological mechanisms underlying complex traits. All variants in the major histocompatibility complex (MHC) region were excluded before running the MAGMA analyses.

Gene mapping is a crucial process for understanding the relationship between genetic variants and traits or diseases. To map the significant SNPs to genes, we used variant to gene (v2g) from Open Targets Genetics^62^, which provides a comprehensive and systematic approach to prioritize causal variants and identify likely causal genes associated with various phenotypes and diseases. By integrating positional data on the distance between the variant and each gene’s canonical transcription start site, eQTL, pQTL, splicingQTL, epigenomic data, and functional prediction, Open Targets maps lead SNPs to genes. We also conducted Gene Ontology gene-set analysis based on FUMA’s gene ontology classification system^58, 63^, and pathway analysis using Consensus PathDB^64^.

### Cell specificity, protein–protein interaction and gene-drug interaction analyses

To investigate the context-specific expression of mapped genes associated with basal ganglia we used WebCSEA (Web-based Cell-type-Specific Enrichment Analysis of Genes). WebCSEA incorporates a curated collection of 111 scRNA-seq panels from various human tissues and 1,355 tissue-cell types across 11 organ systems^65^. We also determined the cell-specific expression within the human cerebral cortex including neurons, fetal and mature astrocytes, oligodendrocytes, microglia/macrophages, and endothelial cells for each mapped gene separately^66^.

In addition, we employed GENEmania^66^ for functional genomics analysis, to investigate protein-protein interactions (PPIs) between protein-encoded basal ganglia-linked genes and exported the network to Cytoscape 3.10. We identified potential protein clusters and gain insights into the functional relationships between the proteins of interest. Finally, we performed gene-drug interaction analysis using the Drug Gene Interaction Database to gain insights into the potential associations between mapped genes to basal ganglia and drugs, including known drug targets and potential off-target effects^67^.

### Genetic overlap between the basal ganglia and common brain disorders

We studied the genetic overlap between basal ganglia and ASD, ADHD, SCZ, BIP, MDD, MIG, PD, and Alz. We used GWAS summary statistics from the Psychiatric Genomics Consortium for ADHD^68^, ASD^69^, BIP^70^, MDD^71^, SCZ^72^ and Alz^73^, from International headache genetics Consortium for MIG ^74^, and from the International Parkinson Disease Genomics Consortium^75, 76^ for PD data. All the studies included in the original GWAS were approved by local ethics authorities.

First, we used linkage disequilibrium (LD) score regression^55, 56^ to estimate genetic correlations between basal ganglia volumes and each of the brain disorders. Then we created conditional quantile-quantile (Q-Q) plots to evaluate the presence of cross-phenotype polygenic enrichment^77, 78^, conditioning basal ganglia on each brain disorder and vice versa. The conditional FDR method ^77–79^ builds on an empirical Bayesian statistical framework and leverages cross-phenotype enrichment observed on the conditional Q-Q plots to improve the discovery of genetic loci associated with the traits of interest. Next, we used the conjunctional FDR (conjFDR) approach at FDR 0.05 ^77–79^ to identify common genetic loci between basal ganglia and the included brain disorders. This approach takes the highest of the two condFDR statistics for a particular SNP, i.e. for trait A conditional on trait B and trait B conditional on trait A. If the p values for both phenotypes are equal to or less than the p values for each trait separately, this reflects an estimate of the posterior probability that a SNP is null for one or both characteristics. More details are found in the original^77, 79^ and subsequent publication^78^. The conjFDR analysis does incorporate effect directions, in contrast to genetic correlation analysis, and may thus be used to analyze summary data from multivariate GWAS that lack effect directions. Because complex correlations in regions with intricate LD can bias FDR estimation, two genomic regions—the extended major histocompatibility complex genes region (hg19 location Chr 6: 25119106-33854733) and chromosome 8p23.1 (hg19 location Chr 8: 7242715-12483982) for all phenotypes and MAPT region for PD and APOE region for Alz and ASD, respectively—were excluded from the FDR-fitting procedures. We used the FUMA protocol to annotate independent genomic loci jointly associated with basal ganglia and each brain disorder. We submitted the results of the conjFDR analysis to FUMA v1.5.1^58^. The lead SNPs and candidate SNPs within each genomic locus were used to identify the genomic loci as significant SNPs with an LD r^2^ >= 0.6 and conjFDR 0.1 and at least one associated independent significant SNP. ConjFDR <0.05 and r^2^ <0.6 were used to determine independent significant SNPs, while lead SNPs were defined if they were in approximate linkage equilibrium with each other (r^2^ < 0.1). Finally, the significant SNPs were mapped to genes using Open Targets^62^ as described above.

## Code Availability

All code and software needed to generate the results is available as part of public resources, specifically MOSTest (https://github.com/precimed/mostest), FUMA (https://fuma.ctglab.nl/), conjunctional FDR (https://github.com/precimed/pleiofdr/) and LD score regression (https://github.com/bulik/ldsc).

## Supplementary Information

**Supplementary Figure 1.**
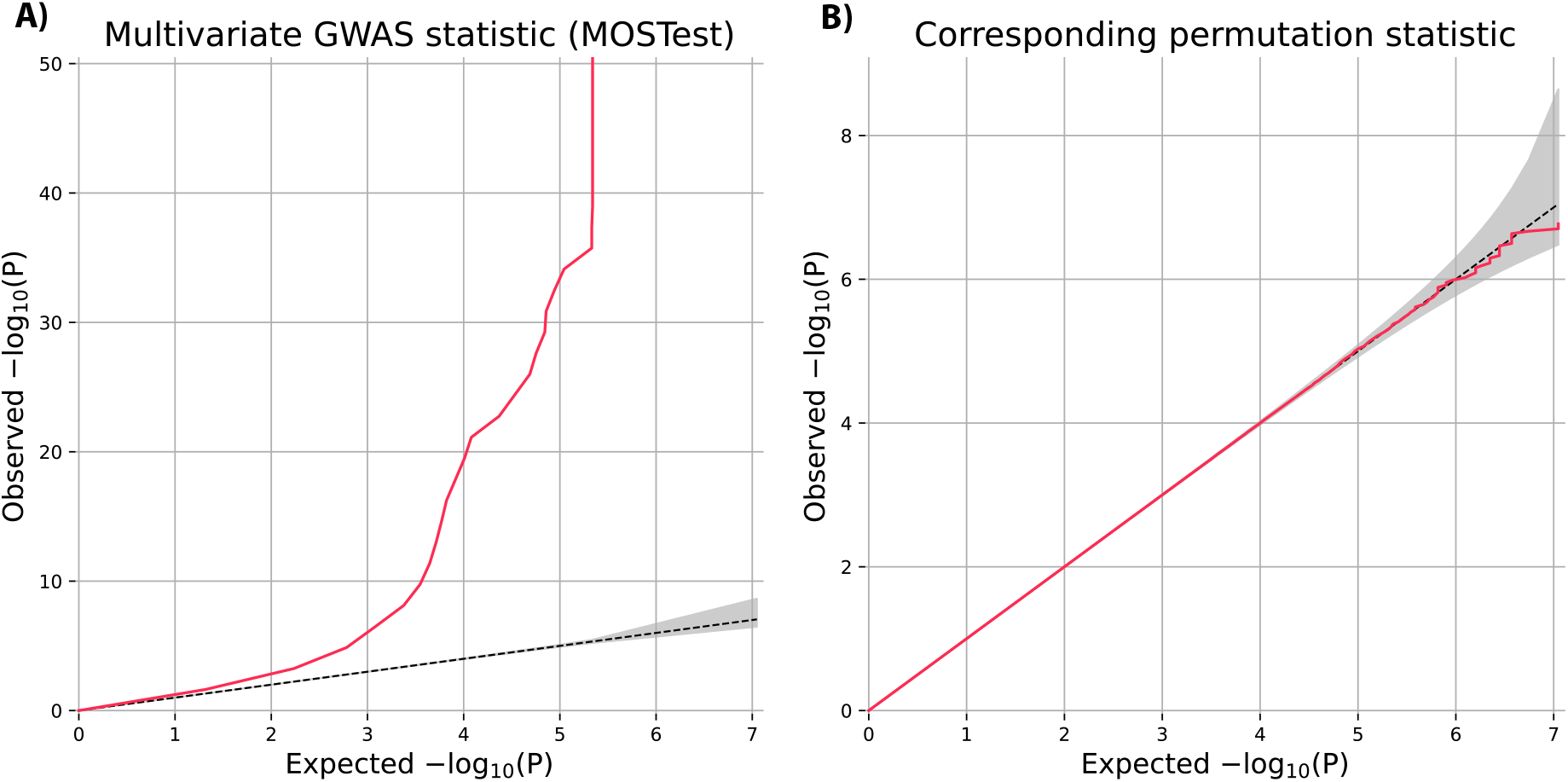
Quantile-quantile plots from MOSTest analysis. **A)** The left panel shows signal from MOSTest analysis. **B)** The right panel (from permutation testing) shows test statistics under null and confirms validity of the MOSTest test statistics.

**Supplementary Figure 2.**
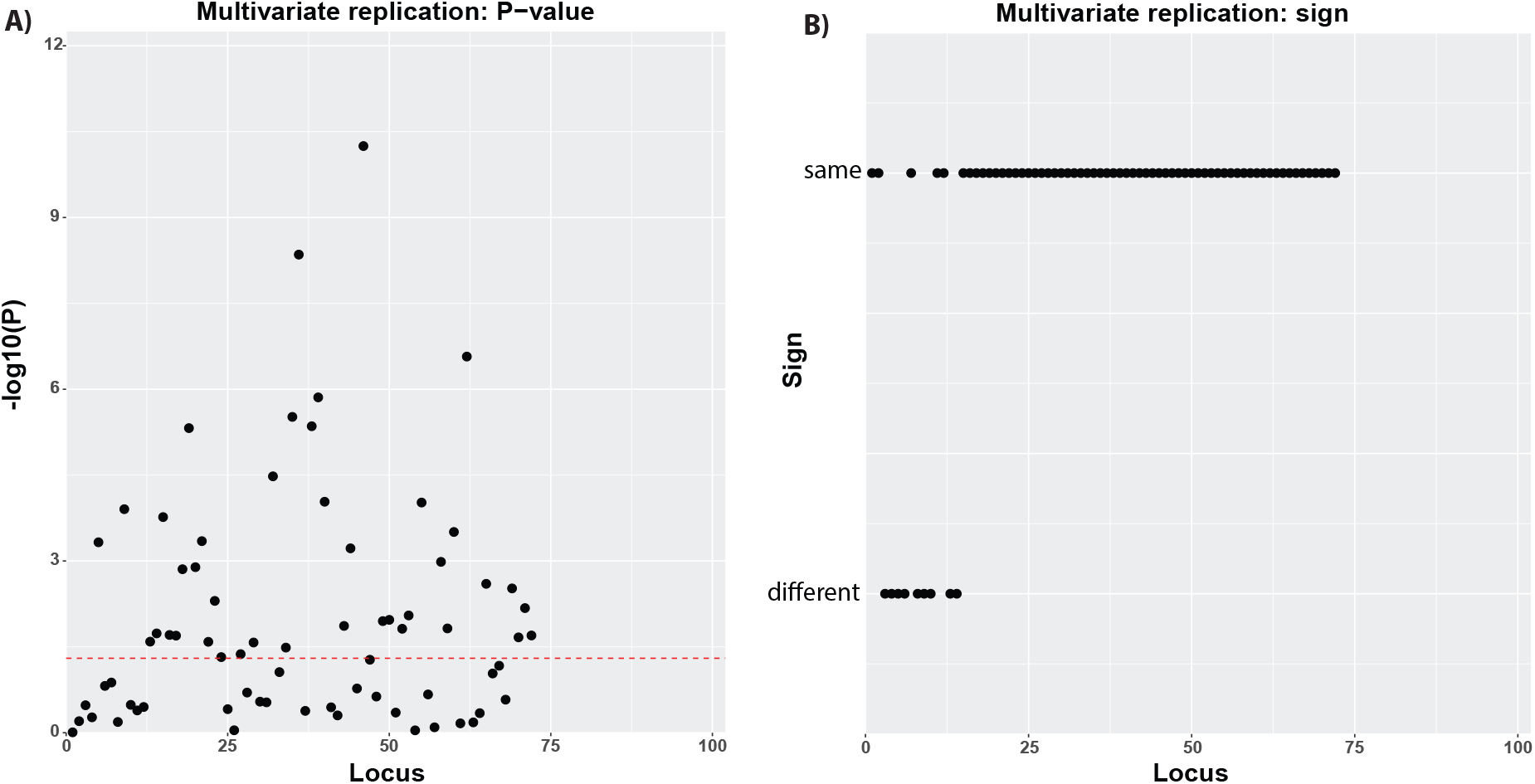
Multivariate replication analysis using independent data from 5220 individuals with non-white ethnicity. Using a multivariate replication procedure (see Methods), we found that 55.6% of the loci replicated at P <0.05 and 87.5% showed the same direction. P-values are denoted as -log_10_(P).

**Supplementary Figure 3.**
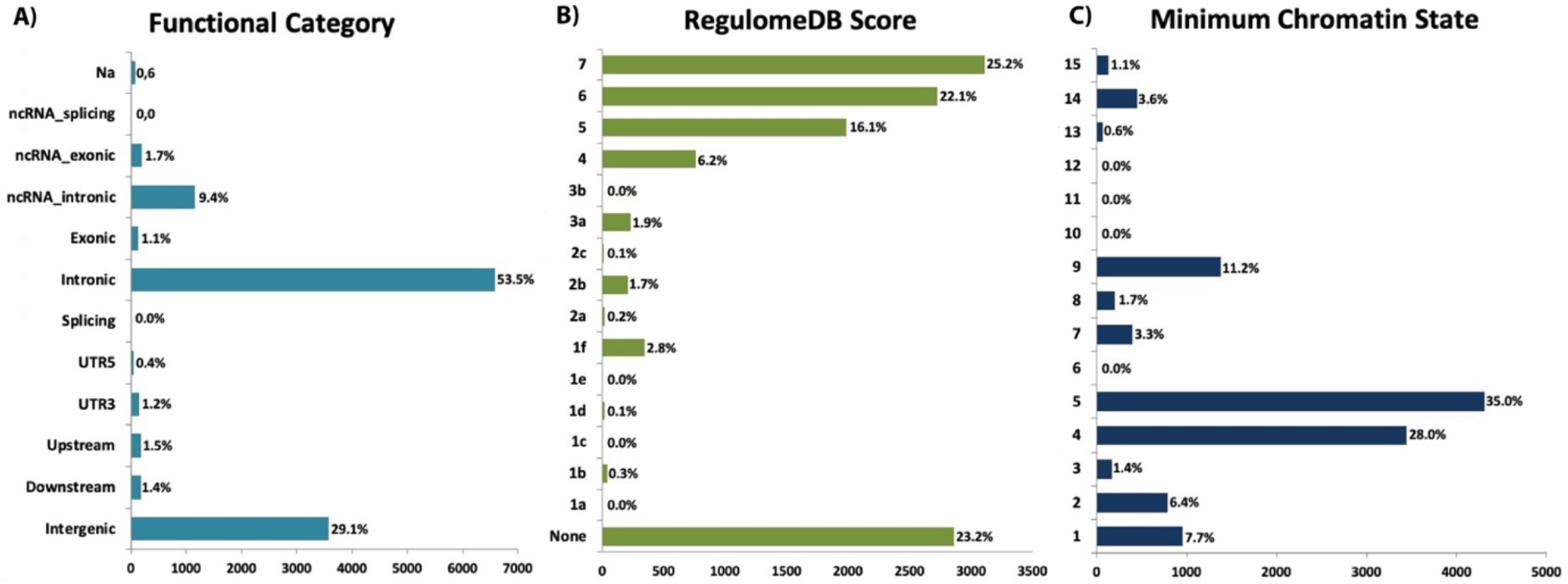
Distribution of the annotation for all SNPs in the significant genetic loci from the basal ganglia GWAS including. (A) the minimum chromatin state across 127 tissue and cell types for SNPs in the significant genomic loci, with lower states indicating higher accessibility and states 1–7 referring to open chromatin states, (B) the distribution of RegulomeDB scores for SNPs in the significant genomic loci, with a low score indicating a higher likelihood of having a regulatory function and (C) the distribution of functional consequences of SNPs in the significant genomic risk loci. The chromatin states are 1=Active Transcription Start Site (TSS); 2=Flanking Active TSS; 3=Transcription at gene 5’ and 3’; 4=Strong transcription; 5=Weak Transcription; 6=Genic enhancers; 7=Enhancers; 8=Zinc finger genes & repeats; 9=Heterochromatic; 10=Bivalent/Poised TSS; 11=Flanking Bivalent/Poised TSS/Enh; 12=Bivalent Enhancer; 13=Repressed PolyComb; 14=Weak Repressed PolyComb; 15=Quiescent/Low. RegulomeDB categories reflect: 1a: eQTL + TF binding + matched TF motif + matched DNase Footprint + DNase peak; 1b: eQTL + TF binding + any motif + DNase Footprint + DNase peak; 1c: eQTL + TF binding + matched TF motif + DNase peak; 1d: eQTL + TF binding + any motif + DNase peak; 1e: eQTL + TF binding + matched TF motif; 1f: eQTL + TF binding / DNase peak; 2a: TF binding + matched TF motif + matched DNase Footprint + DNase peak; 2b: TF binding + any motif + DNase Footprint + DNase peak; 2c: TF binding + matched TF motif + DNase peak; 3a: TF binding + any motif + DNase peak; 3b: TF binding + matched TF motif; 4: TF binding + DNase peak; 5: TF binding or DNase peak; 6: Motif hit; 7: Other.

**Supplementary Figure 4.**
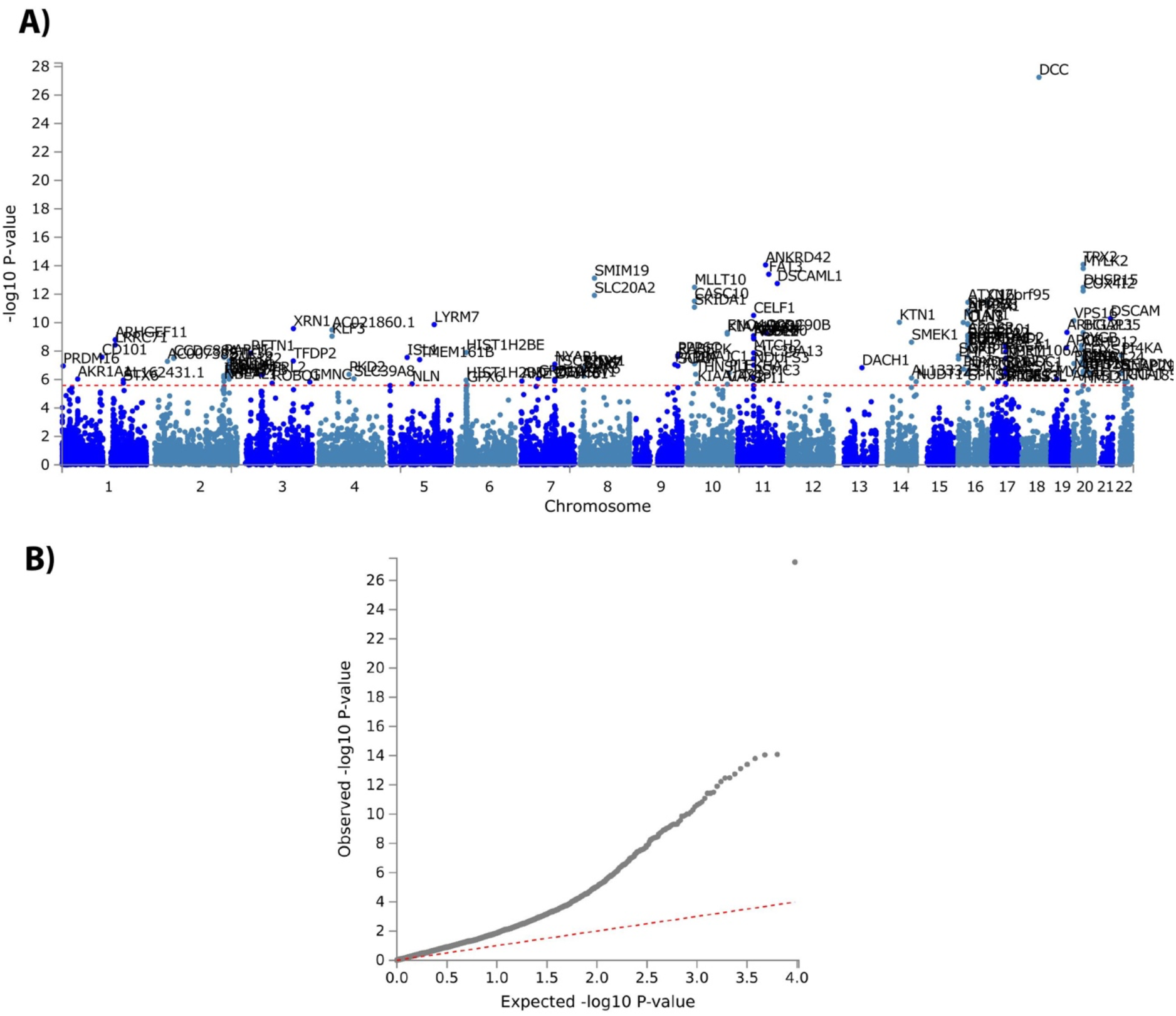
**A.** Manhattan plots from the genome-wide gene-based association analyses for basal ganglia. 149 genes were associated with basal ganglia. The red horizontal lines indicate significance threshold of two-sided P = 2.622e-6. **B.** That is a Q-Q plot of the gene-based test computed by MAGMA.

**Supplementary Figure 5.**
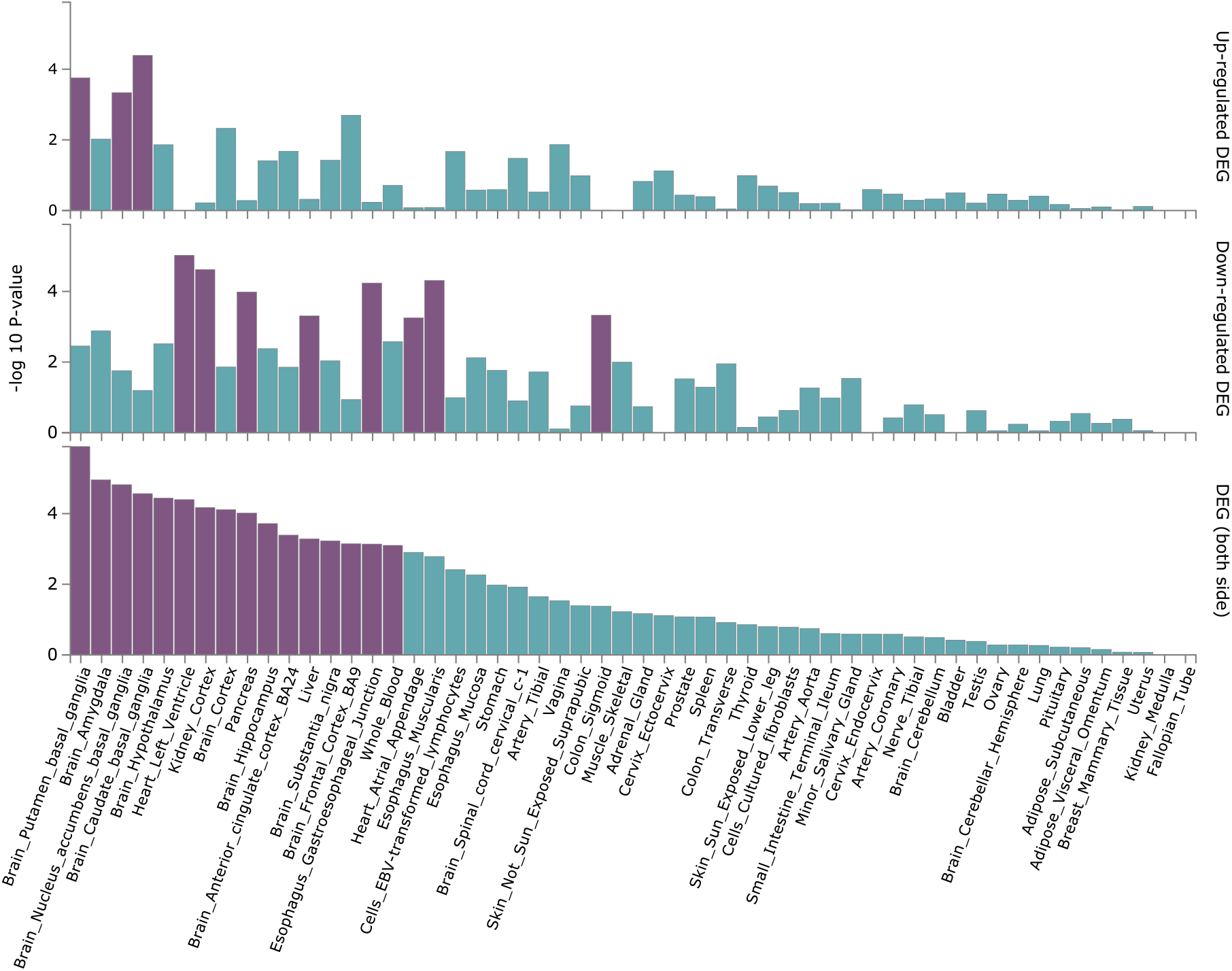
GTEx enrichment analysis based on the 75 genes mapped by Open target. General tissue types.

**Supplementary Figure 6.**
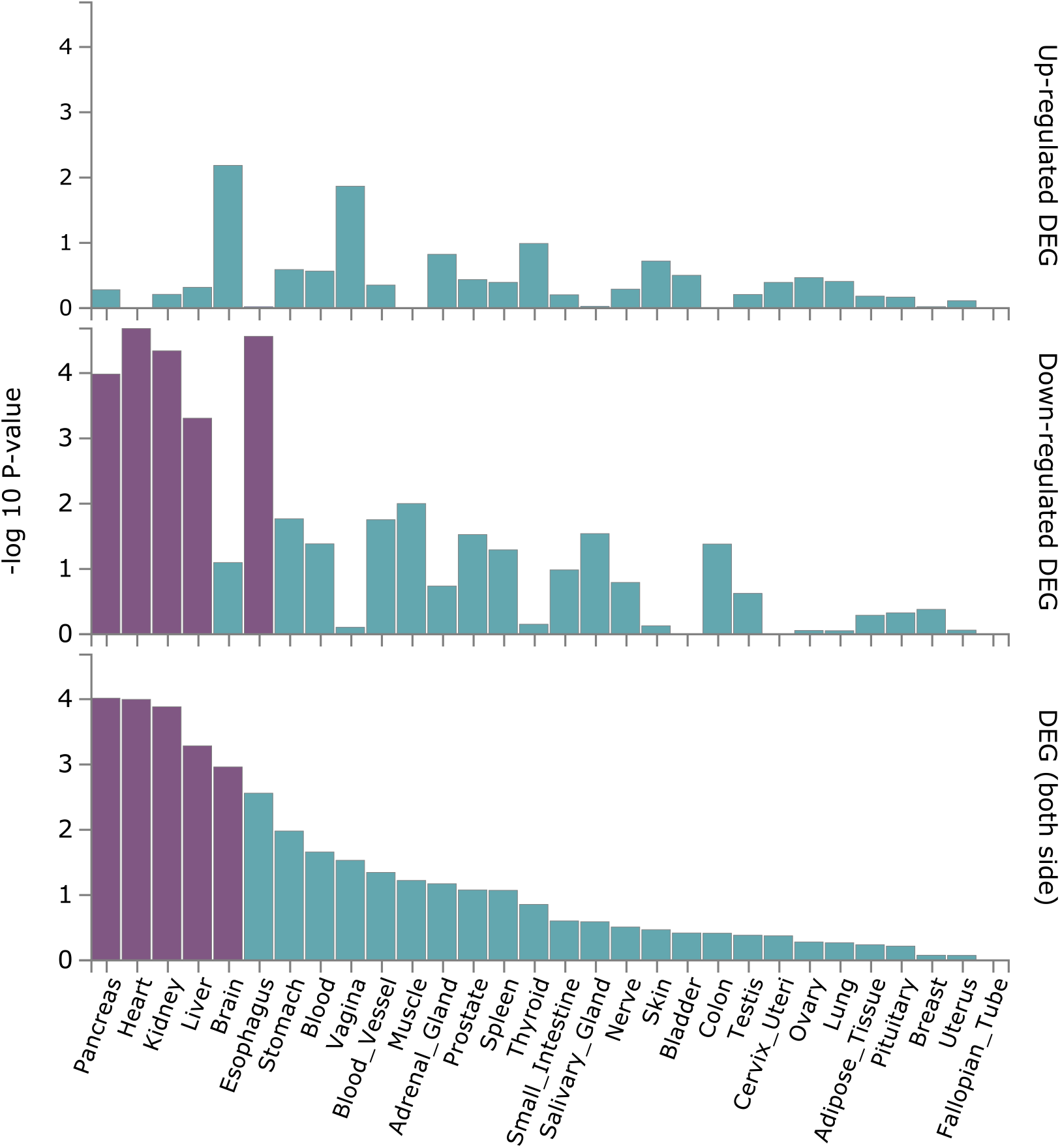
GTEx enrichment analysis based on the 75 genes mapped by Open target. Tissue types. P-values are two-tailed.

**Supplementary Figure 7.**
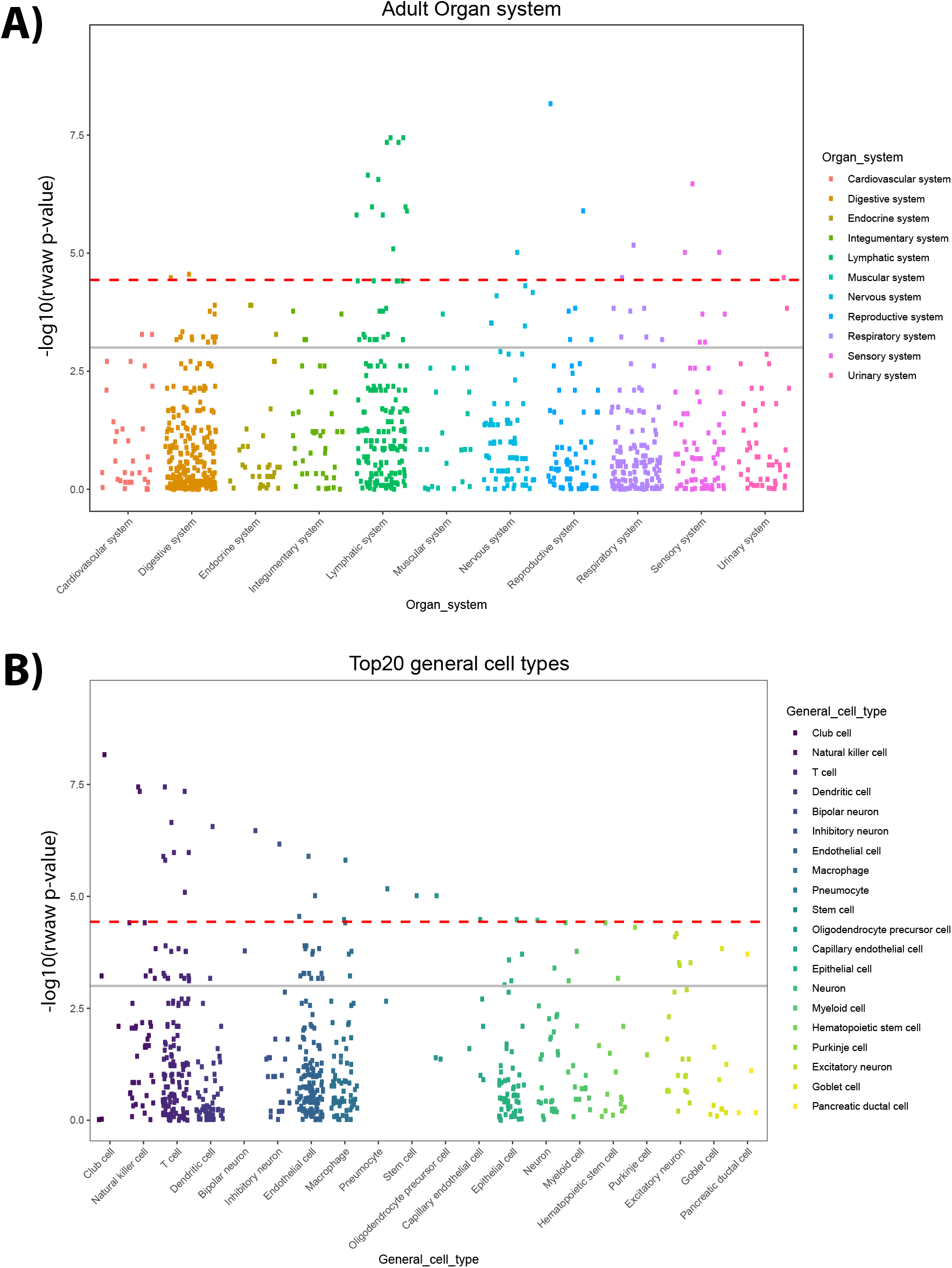
**A.** WebCSEA top enriched adult organ systems. **B.** WebCSEA top enriched cell-types.

**Supplementary Figure 8.**
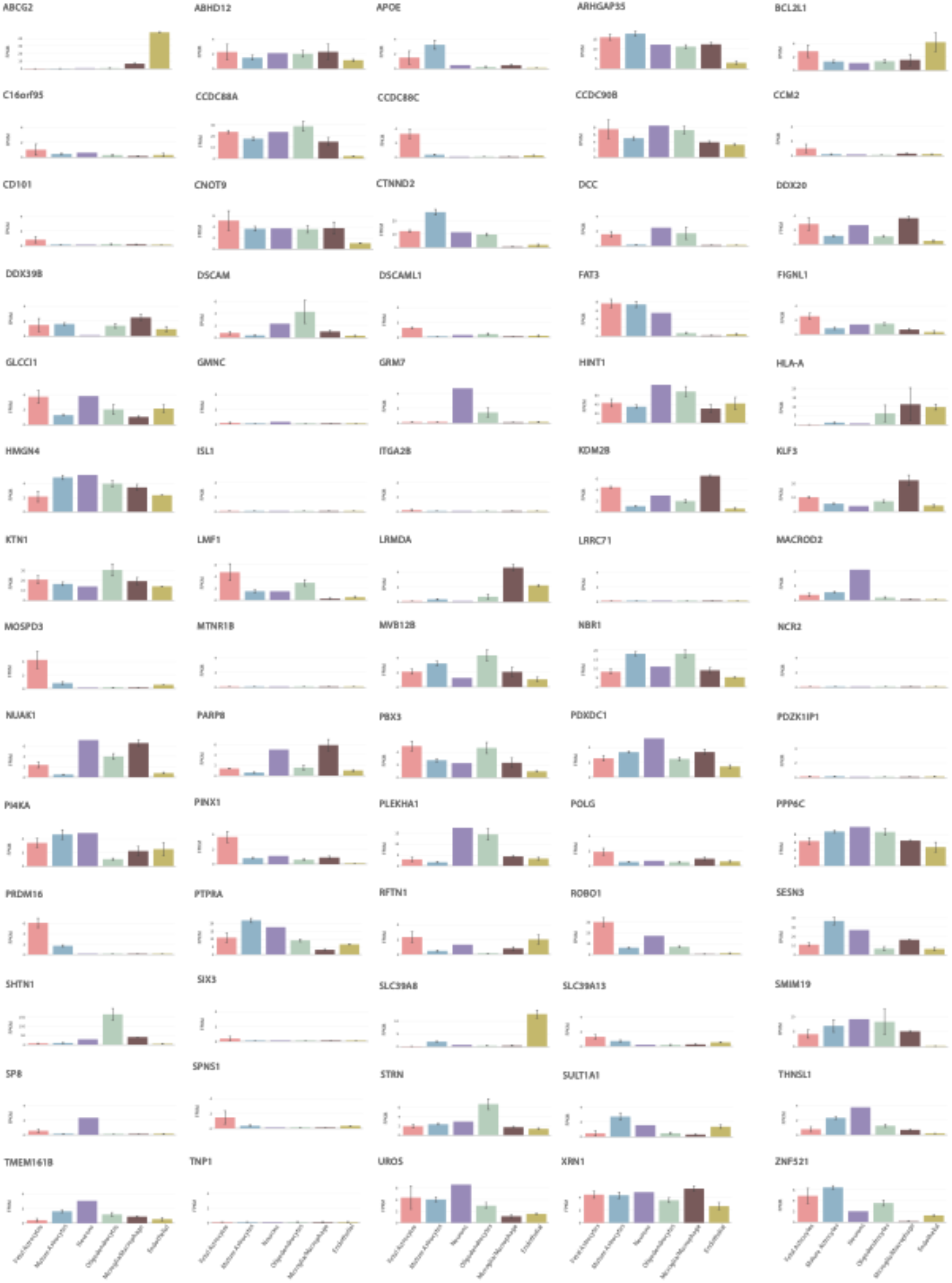
Cell type analysis for the 73 mapped genes. Genes that were not expressed or that weremissing in the data base were not included yielding 70 genes.. Panels shows profiles per gene. FPKM=Fragments Per Kilobase Million.

**Supplementary Figure 9.**
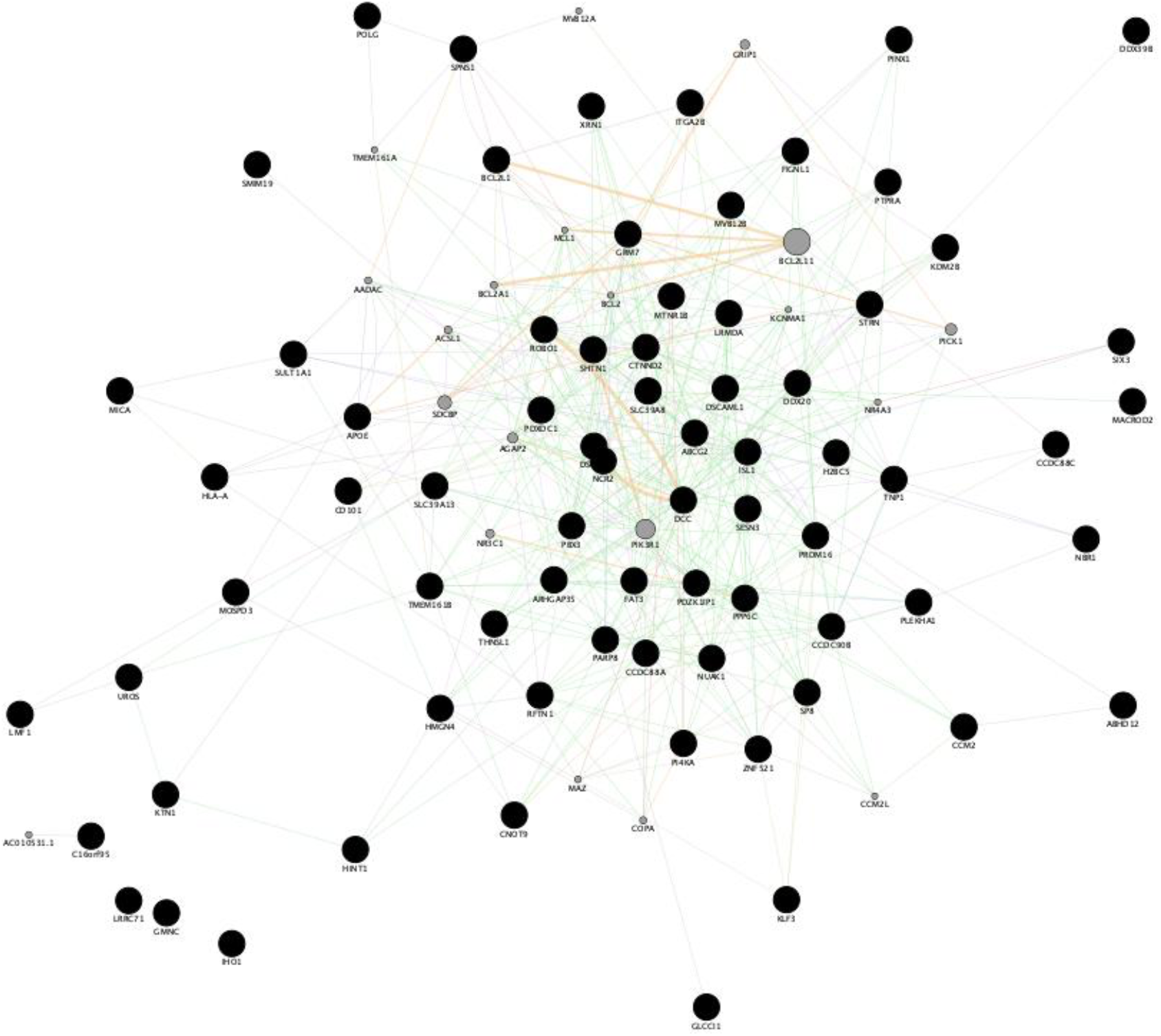
Network interaction graph predominantly illustrating co-expression and shared protein domains for the 74 mapped genes.

**Supplementary Figure 10.**
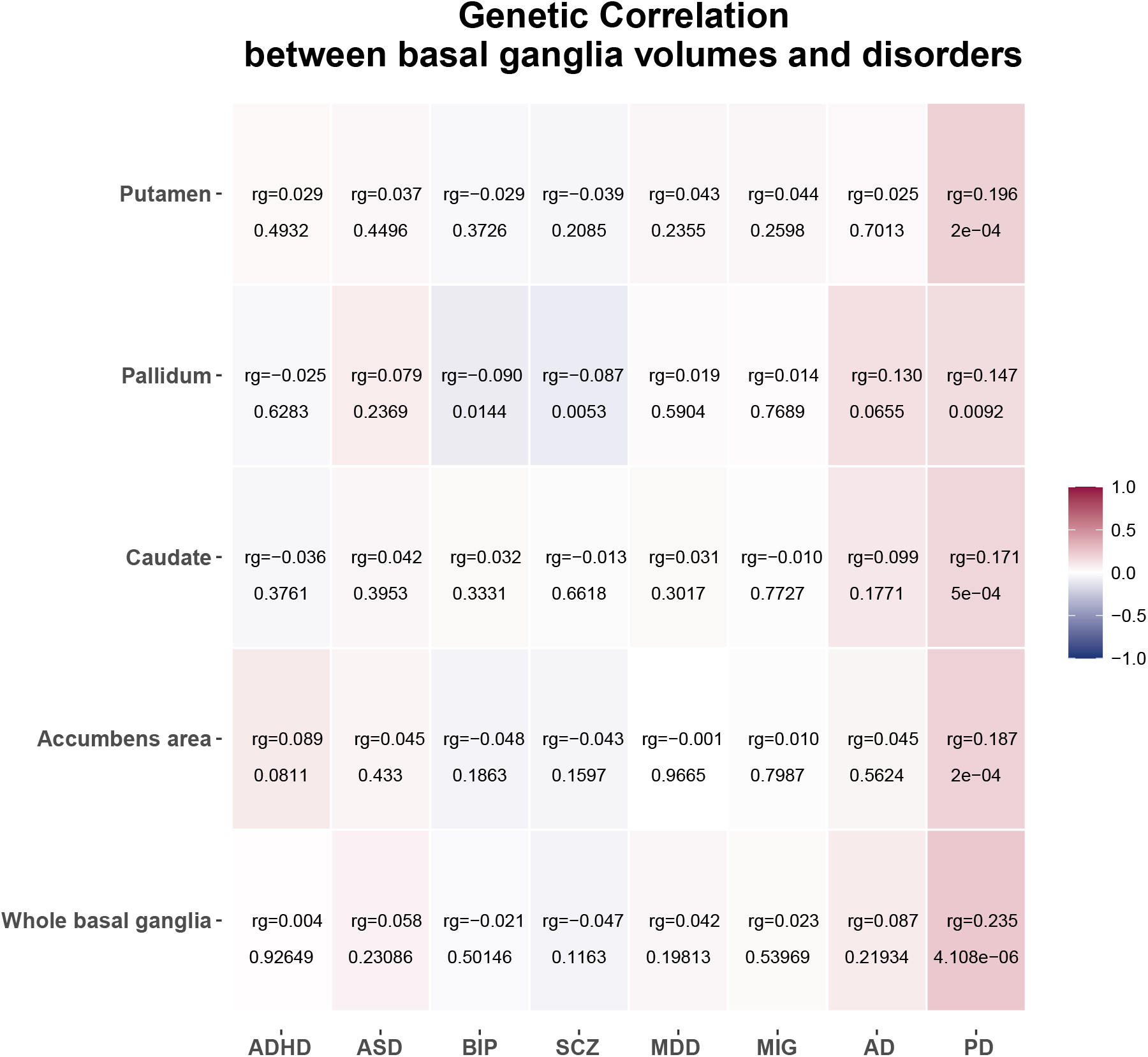
LD-score regression based genetic correlations between basal ganglia volumes and eight brain disorders. The analysis is based on the univariate statistics of the individual regions. Colors reflect correlation strengths. P-values are two-tailed. ADHD; attention-deficit hyperactivity disorder. ASD, autism spectrum disorder; BIP, bipolar disorder; SCZ, schizophrenia; MDD, major depression; MIG: migraine; Alz; Alzheimer’s disease; PD, Parkinson’s disease.

**Supplementary Figure 11.**
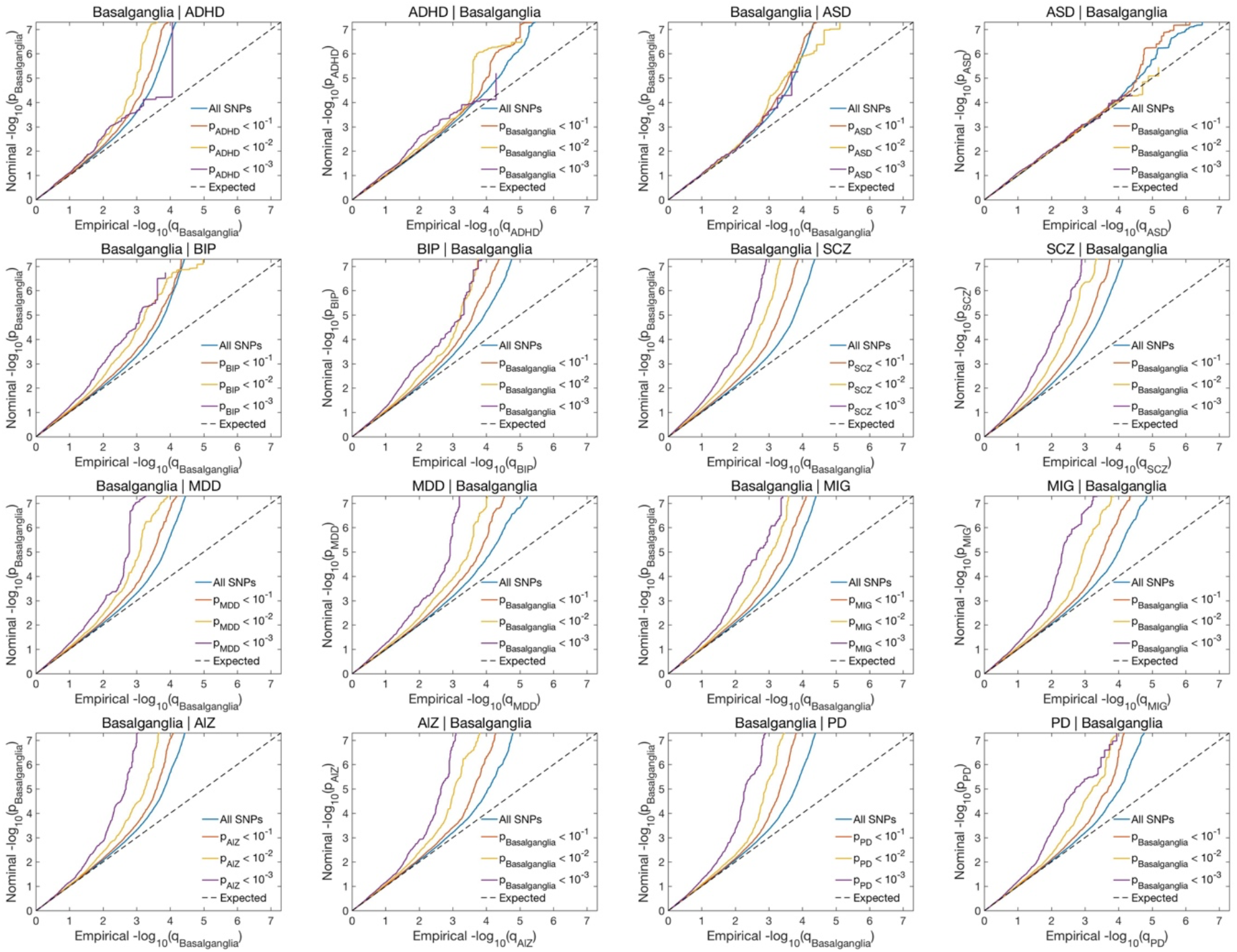
Conditional Q-Q plots for basal ganglia given associations with the disorder (left figures) and vice versa (right figures). ADHD; attention-deficit hyperactivity disorder. ASD, autism spectrum disorder; BIP, bipolar disorder; SCZ, schizophrenia; MDD, major depression; MIG: migraine; AD; Alzheimer’s disease; PD, Parkinson’s disease.

## References

1 Alexander, G. E. & Crutcher, M. D. Functional architecture of basal ganglia circuits: neural substrates of parallel processing. Trends in neurosciences 13, 266–271 (1990).

2 Parent, A. Extrinsic connections of the basal ganglia. Trends in neurosciences 13, 254–258 (1990).

3 Haber, S. N. & Knutson, B. The reward circuit: linking primate anatomy and human imaging. Neuropsychopharmacology: official publication of the American College of Neuropsychopharmacology 35, 4–26 (2010).

4 DeLong, M. R. & Wichmann, T. Circuits and circuit disorders of the basal ganglia. Archives of neurology 64, 20–24 (2007).

5 Grahn, J. A., Parkinson, J. A. & Owen, A. M. The cognitive functions of the caudate nucleus. Progress in neurobiology 86, 141–155 (2008).

6 Delmonte, S., Gallagher, L., O’hanlon, E., McGrath, J. & Balsters, J. H. Functional and structural connectivity of frontostriatal circuitry in Autism Spectrum Disorder. Frontiers in human neuroscience 7, 430 (2013).

7 Redgrave, P. et al. Goal-directed and habitual control in the basal ganglia: implications for Parkinson’s disease. Nature Reviews Neuroscience 11, 760–772 (2010).

8 Yin, H. H. & Knowlton, B. J. The role of the basal ganglia in habit formation. Nature Reviews Neuroscience 7, 464–476 (2006).

9 Graybiel, A. M. Habits, rituals, and the evaluative brain. Annu. Rev. Neurosci. 31, 359–387 (2008).

10 Frank, M. J. Dynamic dopamine modulation in the basal ganglia: a neurocomputational account of cognitive deficits in medicated and nonmedicated Parkinsonism. Journal of cognitive neuroscience 17, 51–72 (2005).

11 D’Cruz, A.-M. et al. Reduced behavioral flexibility in autism spectrum disorders. Neuropsychology 27, 152 (2013).

12 McAlonan, G. M. et al. Brain anatomy and sensorimotor gating in Asperger’s syndrome. Brain: a journal of neurology 125, 1594–1606 (2002).

13 O’doherty, J. P. Reward representations and reward-related learning in the human brain: insights from neuroimaging. Current opinion in neurobiology 14, 769–776 (2004).

14 Schultz, W. Behavioral theories and the neurophysiology of reward. Annu. Rev. Psychol. 57, 87–115 (2006).

15 Haber, S. N. Corticostriatal circuitry. Dialogues in clinical neuroscience (2022).

16 Cogswell, P. M. et al. Associations of quantitative susceptibility mapping with Alzheimer’s disease clinical and imaging markers. Neuroimage 224, 117433 (2021).

17 Mostofsky, S. H. & Ewen, J. B. Altered connectivity and action model formation in autism is autism. The Neuroscientist 17, 437–448 (2011).

18 Langen, M. et al. Changes in the development of striatum are involved in repetitive behavior in autism. Biological psychiatry 76, 405–411 (2014).

19 Mostofsky, S. H. & Simmonds, D. J. Response inhibition and response selection: two sides of the same coin. Journal of cognitive neuroscience 20, 751–761 (2008).

20 Castellanos, F. X., Sonuga-Barke, E. J., Milham, M. P. & Tannock, R. Characterizing cognition in ADHD: beyond executive dysfunction. Trends in cognitive sciences 10, 117–123 (2006).

21 Luman, M., Tripp, G. & Scheres, A. Identifying the neurobiology of altered reinforcement sensitivity in ADHD: a review and research agenda. Neuroscience & Biobehavioral Reviews 34, 744–754 (2010).

22 Volkow, N. D. et al. Motivation deficit in ADHD is associated with dysfunction of the dopamine reward pathway. Molecular psychiatry 16, 1147–1154 (2011).

23 Plichta, M. M. et al. Neural hyporesponsiveness and hyperresponsiveness during immediate and delayed reward processing in adult attention-deficit/hyperactivity disorder. Biological psychiatry 65, 7–14 (2009).

24 Cubillo, A., Halari, R., Smith, A., Taylor, E. & Rubia, K. A review of fronto-striatal and fronto-cortical brain abnormalities in children and adults with Attention Deficit Hyperactivity Disorder (ADHD) and new evidence for dysfunction in adults with ADHD during motivation and attention. cortex 48, 194–215 (2012).

25 Nigg, J. T. Neuropsychologic theory and findings in attention-deficit/hyperactivity disorder: the state of the field and salient challenges for the coming decade. Biological psychiatry 57, 1424–1435 (2005).

26 Sonuga-Barke, E. J. & Fairchild, G. Neuroeconomics of attention-deficit/hyperactivity disorder: differential influences of medial, dorsal, and ventral prefrontal brain networks on suboptimal decision making? Biological psychiatry 72, 126–133 (2012).

27 Solanto, M. V. Dopamine dysfunction in AD/HD: integrating clinical and basic neuroscience research. Behavioural brain research 130, 65–71 (2002).

28 Swanson, J. M. et al. Etiologic subtypes of attention-deficit/hyperactivity disorder: brain imaging, molecular genetic and environmental factors and the dopamine hypothesis. Neuropsychology review 17, 39–59 (2007).

29 Castellanos, F. X. & Tannock, R. Neuroscience of attention-deficit/hyperactivity disorder: the search for endophenotypes. Nature Reviews Neuroscience 3, 617–628 (2002).

30 Howes, O. D. & Kapur, S. The dopamine hypothesis of schizophrenia: version III—the final common pathway. Schizophrenia bulletin 35, 549–562 (2009).

31 Abi-Dargham, A. & Moore, H. Prefrontal DA transmission at D1 receptors and the pathology of schizophrenia. The Neuroscientist 9, 404–416 (2003).

32 Fornito, A., Yoon, J., Zalesky, A., Bullmore, E. T. & Carter, C. S. General and specific functional connectivity disturbances in first-episode schizophrenia during cognitive control performance. Biological psychiatry 70, 64–72 (2011).

33 Andreasen, N. C. et al. Progressive brain change in schizophrenia: a prospective longitudinal study of first-episode schizophrenia. Biological psychiatry 70, 672–679 (2011).

34 Russo, S. J. & Nestler, E. J. The brain reward circuitry in mood disorders. Nature Reviews Neuroscience 14, 609–625 (2013).

35 Phillips, M. L., Ladouceur, C. D. & Drevets, W. C. A neural model of voluntary and automatic emotion regulation: implications for understanding the pathophysiology and neurodevelopment of bipolar disorder. Molecular psychiatry 13, 833–857 (2008).

36 Pizzagalli, D. A. Depression, stress, and anhedonia: toward a synthesis and integrated model. Annual review of clinical psychology 10, 393–423 (2014).

37 Magon, S. et al. Morphological abnormalities of thalamic subnuclei in migraine: a multicenter MRI study at 3 tesla. Journal of Neuroscience 35, 13800–13806 (2015).

38 Lewis, M. M. et al. The pattern of gray matter atrophy in Parkinson’s disease differs in cortical and subcortical regions. Journal of neurology 263, 68–75 (2016).

39 Beyer, J. L. et al. Caudate volume measurement in older adults with bipolar disorder. International journal of geriatric psychiatry 19, 109–114 (2004).

40 Ellison-Wright, I., Ellison-Wright, Z. & Bullmore, E. Structural brain change in attention deficit hyperactivity disorder identified by meta-analysis. BMC psychiatry 8, 1–8 (2008).

41 Nakao, T., Radua, J., Rubia, K. & Mataix-Cols, D. Gray matter volume abnormalities in ADHD: voxel-based meta-analysis exploring the effects of age and stimulant medication. American Journal of Psychiatry 168, 1154–1163 (2011).

42 Kim, J. et al. Regional grey matter changes in patients with migraine: a voxel-based morphometry study. Cephalalgia 28, 598–604 (2008).

43 Rojas, D. C. et al. Hippocampus and amygdala volumes in parents of children with autistic disorder. American Journal of Psychiatry 161, 2038–2044 (2004).

44 Strakowski, S. M. et al. Ventricular and periventricular structural volumes in first-versus multiple-episode bipolar disorder. American Journal of Psychiatry 159, 1841–1847 (2002).

45 DelBello, M. P., Zimmerman, M. E., Mills, N. P., Getz, G. E. & Strakowski, S. M. Magnetic resonance imaging analysis of amygdala and other subcortical brain regions in adolescents with bipolar disorder. Bipolar disorders 6, 43–52 (2004).

46 Wilke, M., Kowatch, R. A., DelBello, M. P., Mills, N. P. & Holland, S. K. Voxel-based morphometry in adolescents with bipolar disorder: first results. Psychiatry Research: Neuroimaging 131, 57–69 (2004).

47 Estes, A. et al. Basal ganglia morphometry and repetitive behavior in young children with autism spectrum disorder. Autism Research 4, 212–220 (2011).

48 Sears, L. L., et al. An MRI study of the basal ganglia in autism. Progress in neuro-psychopharmacology & biological psychiatry (1999).

49 Hibar, D. P. et al. Common genetic variants influence human subcortical brain structures. Nature 520, 224–229 (2015).

50 Satizabal, C. L. et al. Genetic architecture of subcortical brain structures in 38,851 individuals. Nature genetics 51, 1624–1636 (2019).

51 Bycroft, C. et al. The UK Biobank resource with deep phenotyping and genomic data. Nature 562, 203–209 (2018). https://doi.org:10.1038/s41586-018-0579-z

52 Fischl, B. et al. Whole brain segmentation: automated labeling of neuroanatomical structures in the human brain. Neuron 33, 341–355 (2002).

53 Fortin, J. P. et al. Harmonization of cortical thickness measurements across scanners and sites. Neuroimage 167, 104–120 (2018). https://doi.org:10.1016/j.neuroimage.2017.11.024

54 van der Meer, D. et al. Understanding the genetic determinants of the brain with MOSTest. Nature communications 11, 1–9 (2020).

55 Bulik-Sullivan, B. K. et al. LD Score regression distinguishes confounding from polygenicity in genome-wide association studies. Nature genetics 47, 291–295 (2015).

56 Bulik-Sullivan, B. et al. An atlas of genetic correlations across human diseases and traits. Nature genetics 47, 1236 (2015).

57 Loughnan, R. J. et al. Generalization of cortical MOSTest genome-wide associations within and across samples. Neuroimage 263, 119632 (2022). https://doi.org:10.1016/j.neuroimage.2022.119632

58 Watanabe, K., Taskesen, E., Van Bochoven, A. & Posthuma, D. Functional mapping and annotation of genetic associations with FUMA. Nature communications 8, 1–11 (2017).

59 Kircher, M. et al. A general framework for estimating the relative pathogenicity of human genetic variants. Nature genetics 46, 310–315 (2014).

60 Boyle, A. P. et al. Annotation of functional variation in personal genomes using RegulomeDB. Genome research 22, 1790–1797 (2012).

61 de Leeuw, C. A., Mooij, J. M., Heskes, T. & Posthuma, D. MAGMA: generalized gene-set analysis of GWAS data. PLoS Comput Biol 11, e1004219 (2015). https://doi.org:10.1371/journal.pcbi.1004219

62 Ochoa, D. et al. The next-generation Open Targets Platform: reimagined, redesigned, rebuilt. Nucleic acids research 51, D1353–d1359 (2023). https://doi.org:10.1093/nar/gkac1046

63 Consortium, G. O. The Gene Ontology resource: enriching a GOld mine. Nucleic acids research 49, D325–d334 (2021). https://doi.org:10.1093/nar/gkaa1113

64 Herwig, R., Hardt, C., Lienhard, M. & Kamburov, A. Analyzing and interpreting genome data at the network level with ConsensusPathDB. Nature protocols 11, 1889–1907 (2016).

65 Dai, Y. et al. WebCSEA: web-based cell-type-specific enrichment analysis of genes. Nucleic acids research 50, W782–w790 (2022). https://doi.org:10.1093/nar/gkac392

66 Franz, M. et al. GeneMANIA update 2018. Nucleic acids research 46, W60–w64 (2018). https://doi.org:10.1093/nar/gky311

67 Freshour, S. L. et al. Integration of the Drug-Gene Interaction Database (DGIdb 4.0) with open crowdsource efforts. Nucleic acids research 49, D1144–d1151 (2021). https://doi.org:10.1093/nar/gkaa1084

68 Demontis, D. et al. Discovery of the first genome-wide significant risk loci for attention deficit/hyperactivity disorder. Nature genetics 51, 63–75 (2019).

69 Grove, J. et al. Identification of common genetic risk variants for autism spectrum disorder. Nature genetics 51, 431–444 (2019). https://doi.org:10.1038/s41588-019-0344-8

70 Mullins, N. et al. Genome-wide association study of more than 40,000 bipolar disorder cases provides new insights into the underlying biology. Nature genetics 53, 817–829 (2021). https://doi.org:10.1038/s41588-021-00857-4

71 Wray, N. R. et al. Genome-wide association analyses identify 44 risk variants and refine the genetic architecture of major depression. Nature genetics 50, 668–681 (2018). https://doi.org:10.1038/s41588-018-0090-3

72 Pardiñas, A. F. et al. Common schizophrenia alleles are enriched in mutation-intolerant genes and in regions under strong background selection. Nature genetics 50, 381–389 (2018). https://doi.org:10.1038/s41588-018-0059-2

73 Wightman, D. P. et al. A genome-wide association study with 1,126,563 individuals identifies new risk loci for Alzheimer’s disease. Nature genetics 53, 1276–1282 (2021).

74 Gormley, P. et al. Meta-analysis of 375,000 individuals identifies 38 susceptibility loci for migraine. Nature genetics 48, 856–866 (2016).

75 Nalls, M. A. et al. Large-scale meta-analysis of genome-wide association data identifies six new risk loci for Parkinson’s disease. Nature genetics 46, 989–993 (2014). https://doi.org:10.1038/ng.3043

76 Nalls, M. A. et al. Identification of novel risk loci, causal insights, and heritable risk for Parkinson’s disease: a meta-analysis of genome-wide association studies. The Lancet Neurology 18, 1091–1102 (2019).

77 Andreassen, O. A. et al. Improved detection of common variants associated with schizophrenia by leveraging pleiotropy with cardiovascular-disease risk factors. The American Journal of Human Genetics 92, 197–209 (2013).

78 Smeland, O. B. et al. Discovery of shared genomic loci using the conditional false discovery rate approach. Human genetics 139, 85–94 (2020).

79 Andreassen, O. A. et al. Improved detection of common variants associated with schizophrenia and bipolar disorder using pleiotropy-informed conditional false discovery rate. PLoS genetics 9, e1003455 (2013).

80 Kundaje, A. et al. Integrative analysis of 111 reference human epigenomes. Nature 518, 317–330 (2015).

81 Smeland, O. B. et al. Genome-wide association analysis of Parkinson’s disease and schizophrenia reveals shared genetic architecture and identifies novel risk loci. Biological psychiatry 89, 227–235 (2021).

82 Pickrell, J. K. et al. Detection and interpretation of shared genetic influences on 42 human traits. Nature genetics 48, 709–717 (2016). https://doi.org:10.1038/ng.3570

83 Leonenko, G. et al. Identifying individuals with high risk of Alzheimer’s disease using polygenic risk scores. Nature communications 12, 4506 (2021). https://doi.org:10.1038/s41467-021-24082-z

84 Kulminski, A. M. et al. Genetic and regulatory architecture of Alzheimer’s disease in the APOE region. *Alzheimer’s & dementia (Amsterdam*, Netherlands) 12, e12008 (2020). https://doi.org:10.1002/dad2.12008

85 Deans, M. R. et al. Control of neuronal morphology by the atypical cadherin Fat3. Neuron 71, 820–832 (2011).

86 Santama, N., Er, C. P., Ong, L.-L. & Yu, H. Distribution and functions of kinectin isoforms. Journal of cell science 117, 4537–4549 (2004).

87 Mu, W., Tochen, L., Bertsch, C., Singer, H. S. & Barañano, K. W. Intracranial calcifications and dystonia associated with a novel deletion of chromosome 8p11.2 encompassing SLC20A2 and THAP1. BMJ Case Rep 12 (2019). https://doi.org:10.1136/bcr-2018-228782

88 Hsu, S. C. et al. Mutations in SLC20A2 are a major cause of familial idiopathic basal ganglia calcification. Neurogenetics 14, 11–22 (2013).

89 Taglia, I., Bonifati, V., Mignarri, A., Dotti, M. T. & Federico, A. Primary familial brain calcification: update on molecular genetics. Neurological Sciences 36, 787–794 (2015).

90 He, L. et al. ZIP8, member of the solute-carrier-39 (SLC39) metal-transporter family: characterization of transporter properties. Molecular pharmacology 70, 171–180 (2006).

91 Choi, E.-K., Nguyen, T.-T., Gupta, N., Iwase, S. & Seo, Y. A. Functional analysis of SLC39A8 mutations and their implications for manganese deficiency and mitochondrial disorders. Sci Rep-Uk 8, 3163 (2018).

92 Horning, K. J., Caito, S. W., Tipps, K. G., Bowman, A. B. & Aschner, M. Manganese is essential for neuronal health. Annual review of nutrition 35, 71–108 (2015).

93 Kong, L. et al. The ubiquitin E3 ligase TRIM10 promotes STING aggregation and activation in the Golgi apparatus. Cell Reports 42 (2023).

94 Tamouza, R., Krishnamoorthy, R. & Leboyer, M. Understanding the genetic contribution of the human leukocyte antigen system to common major psychiatric disorders in a world pandemic context. Brain, behavior, and immunity 91, 731–739 (2021).

95 Endres, D. et al. Immunological causes of obsessive-compulsive disorder: is it time for the concept of an “autoimmune OCD” subtype? Translational psychiatry 12, 5 (2022).

96 Jiang, Q. et al. ApoE promotes the proteolytic degradation of Aβ. Neuron 58, 681–693 (2008).

97 Odagiri, S. et al. Autophagic adapter protein NBR1 is localized in Lewy bodies and glial cytoplasmic inclusions and is involved in aggregate formation in α-synucleinopathy. Acta neuropathologica 124, 173–186 (2012).

98 Lange, S. et al. The kinase domain of titin controls muscle gene expression and protein turnover. Science 308, 1599–1603 (2005).

99 Smith, T. M. et al. Complete genomic sequence and analysis of 117 kb of human DNA containing the gene BRCA1. Genome research 6, 1029–1049 (1996).

100 Trachtenberg, J. T. et al. Long-term in vivo imaging of experience-dependent synaptic plasticity in adult cortex. Nature 420, 788–794 (2002).

101 Kozorovitskiy, Y., Saunders, A., Johnson, C. A., Lowell, B. B. & Sabatini, B. L. Recurrent network activity drives striatal synaptogenesis. Nature 485, 646–650 (2012).

102 Carrascoza, F. & Silaghi-Dumitrescu, R. The dynamics of hemoglobin-haptoglobin complexes. Relevance for oxidative stress. Journal of Molecular Structure 1250, 131703 (2022).

103 Hyde, C. L. et al. Identification of 15 genetic loci associated with risk of major depression in individuals of European descent. Nature genetics 48, 1031–1036 (2016).

104 Yue, S. et al. Gene-gene interaction and new onset of major depressive disorder: Findings from a Chinese freshmen nested case-control study. Journal of affective disorders 300, 505–510 (2022). https://doi.org:10.1016/j.jad.2021.12.138

105 Muench, C. et al. The major depressive disorder GWAS-supported variant rs10514299 in TMEM161B-MEF2C predicts putamen activation during reward processing in alcohol dependence. Translational psychiatry 8, 131 (2018). https://doi.org:10.1038/s41398-018-0184-9

106 Wang, L. et al. TMEM161B modulates radial glial scaffolding in neocortical development. Proceedings of the National Academy of Sciences of the United States of America 120, e2209983120 (2023). https://doi.org:10.1073/pnas.2209983120

